# A subset of pro-inflammatory CXCL10+ LILRB2+ macrophages derives from recipient monocytes and drives renal allograft rejection

**DOI:** 10.1101/2025.04.21.25324260

**Authors:** Alexis Varin, Jovanne Palvair, Lennie Messager, Jamal Bamoulid, Yacine Benchikh, Jasper Callemeyn, Mélanie Chaintreuil, Ludivine Dal Zuffo, Didier Ducloux, Imane Farhat, Mathieu Legendre, Laurent Martin, Florian Renosi, Xavier Roussel, Thibaut Vaulet, Maarten Naesens, Claire Tinel, Baptiste Lamarthée

**Affiliations:** Université Marie et Louis Pasteur, EFS BFC, Inserm UMR Right, Besançon, France; Department of Nephrology, Dialysis and Renal Transplantation, Besançon University Hospital, Besançon, France; Department of Microbiology, Immunology and Transplantation, Nephrology and Kidney Transplantation Research Group, KU Leuven, Leuven, Belgium; Department of Nephrology and Kidney Transplantation, Dijon University Hospital, Université Bourgogne-Europe, Dijon, France; Department of Pathology, Dijon University Hospital, Université Bourgogne-Europe, Dijon, France

**Author notes:** These authors equally contributed and share first authorship (Alexis Varin, Jovanne Palvair, Lennie Messager). These authors equally contributed and share last authorship (Claire Tinel, Baptiste Lamarthée). Corresponding author. Baptiste Lamarthée PhD rue du Dr Girod, Besançon, France.

## Abstract

In solid organ transplantation, monocytes and macrophages play a cross-cutting role in the rejection process, irrespective of the transplanted tissue and the type of rejection. Here, we integrated multiple single-cell assays (>150,000 cells) with a broad spectrum of blood-derived and renal allograft-derived cells. We observed 6 myeloid cell trajectories enriched in the allograft during rejection, ranging from circulating CD14+ monocytes to differentiated macrophages in the kidney, with one trajectory culminating in a pro-inflammatory macrophage expressing *CXCL9* and *CXCL10*. By analyzing over 850 biopsies using deconvolution, we report that they are absent in pre-transplant allografts, while these *CXCL10*+ macrophages are the immune cells most associated with inflammation during acute rejection. Furthermore, a survival study of over 500 biopsies indicates that they increase the risk of graft loss independently of other immune cells. *CXCL10*+ macrophages differentiate from recipient monocytes, and we have identified 6 major genes associated with their differentiation, including *LILRB2*. In vitro, mimicking allogenic activation of blood monocytes via the CD47/SIRP-a axis induced overexpression of LILRB2, suggesting that *CXCL10*+ macrophages are activated by this pathway. Finally, we show that macrophages overexpressing LILRB2 induce the proliferation of autologous T lymphocytes. Altogether, the present study provides further insight into the pro-inflammatory axes of recipient-derived monocytes/macrophages, and suggests LILRB2 as a therapeutic target.

## Introduction

In the field of solid organ transplantation, graft rejection remains one of the major challenges to ensuring the survival and optimal function of the transplanted organ. While this phenomenon is influenced by various cell types and immune mechanisms*^1^*, monocytes and macrophages play a crucial and cross-cutting role in the rejection process, regardless of the transplanted tissue or histological picture of rejection*^2^*. A recent study has notably consolidated a vast amount of transcriptomic data from multiple solid organ transplants, including kidney, heart, liver, and lungs, revealing a common feature in the rejection of these organs: myeloid cell-driven inflammation*^3^*. This study identified the myeloid-derived CXCL9 and CXCL10 chemokines as central players in this inflammatory process, highlighting a shared mechanism of rejection across different transplanted tissues*^4^*. The ability of CXCL9 and CXCL10 chemokines to be induced in pan-organ rejection, is reflected by the fact that they form excellent biomarkers of rejection*^5–8^*, as illustrated by their routine measurement in the urine of kidney transplant patients to monitor immune quiescence*^9^*. Interestingly, this research suggests that current phenotypes of rejection, against a background of T-cell targeted immunosuppression, does not longer appear to be primarily mediated by antibodies or T lymphocytes, but rather by monocytes and macrophages.

Additionally, three decades of animal model research demonstrate that monocytes are not merely passive players in the immune response but are capable of directly recognizing allogeneic tissues*^10^*. These studies have showed that monocytes, independently of the adaptive immune response, can initiate*^11^* and sustain alloimmune rejection*^12^*. This capacity of monocytes and macrophages to act as primary drivers of histological lesions, through mechanisms outside the classic T and B cell responses*^13^* reinforces their central role in the immune response to pan-organ transplants. These findings challenge the prevailing understanding of rejection by the adaptive immune system and emphasize the central role of myeloid cells in the inflammatory response leading to graft failure.

Monocytes and macrophages are particularly involved through their ability to infiltrate the allograft, contributing to the initiation and amplification of the immune response and the progressive degradation of the graft. However, despite their recognized importance, the precise mechanisms by which circulating monocytes, infiltrate the transplanted organ and differentiate into fully functional macrophages within the renal graft remain largely unknown.

Here, we aimed to conduct an in-depth analysis of myeloid cell populations in the context of kidney graft rejection. By leveraging single-cell sequencing technologies, we established a high-resolution cellular profile of over 150,000 cells derived from the blood of 13 patients (including 5 without rejection and 8 with rejection) and from kidney grafts of 33 patients (15 without rejection and 18 with rejection). We identified differentiation trajectories from circulating monocytes to tissue-resident macrophages and we identified key genes such as *FCN1*, *LILRA2*, *LILRB2*, *LILRA5*, *LST1* and *LY6E* involved in the differentiation of proinflammatory macrophages driving graft rejection. Finally, we explored in vitro and in vivo the significance of these molecules in allogeneic myeloid responses.

## Methods

### Single-cell RNA-sequencing data analysis

#### Main object generation

The current study is a retrospective analysis of 46 publicly available single-cell RNASeq datasets from kidney transplanted patients, either peripheral blood mononuclear cells (PBMCs) or allograft-derived cells harvested from kidney biopsies. Raw data were downloaded either from the NCBI’s Gene Expression Omnibus database (accession number: GSE145927*^14^*, GSE171374*^15^* and GSE140989*^16^*), from BioStudies (accession number E-MTAB-11450*^17^* and E-MTAB-12051*^18^*) or from NCBI’s BioProject (accession number: PRJNA974568*^19^*). Count matrices were filtered using the following parameters: cells having <300 and >10,000 genes detected and circulating cells presenting >10% mitochondrial transcripts were excluded whereas biopsy-derived cells presenting >25% mitochondrial transcripts were excluded. Filtered gene expression matrices were then merged into a single object using the Seurat R package version 5.0.1*^20^*, normalized with Seurat’s NormalizeData function as well as scaled and centered using Seurat’s ScaleData function. The top 2,000 most variable genes were calculated with Seurat’s FindVariableFeatures function prior to calculating principal components (PC) using Seurat’s RunPCA function. Integration was performed using rpca method in Seurat’s IntegrateLayers function*^20^*, on the supercomputer facilities of the Mésocentre de Franche-Comté. UMAP coordinates were calculated from the top 50 PC using Seurat’s RunUMAP function and visualized with Seurat’s DimPlot function. Unsupervised clustering was performed using Seurat’s FindNeighbors and FindClusters functions with a resolution of 0.47, which led to the identification of 23 distinct clusters. Cell types were annotated by comparing the most differentially expressed genes (DEG) in each cluster, calculated using Seurat’s FindAllmarkers function, with canonical markers from the litterature*^14–19,21–26^*; some highly specific DEG found in each cluster and previously unreported in the literature were also highlighted alongside canonical markers. Dot plots and cell proportion barplots were generated using respectively the DotPlot_Heatmap and Barplot_Cell_Proportion functions from the R package RightOmicsTools version 2.2.0*^27^*, with normalized counts as input data.

#### Myeloid cells reintegration

Cells identified as monocytes, macrophages and cDC were subsetted from the main object and reintegrated in order to determine their cell subtypes composition. Briefly, the top 2,000 most variable genes were recalculated, prior to a new integration with rpca method. UMAP coordinates and unsupervised clustering were calculated using Seurat’s respective functions from the top 50 PC and a resolution of 0.85, which led to the identification of 11 distinct clusters. Annotation was performed on Seurat’s FindAllMarkers top DEG in each cluster and compared to canonical markers from the literature*^14–19,28,29^*. Seurat’s FindMarkers function was used to find DEG between *SELENOP*+ macrophages and *CXCL10*+ macrophages, and a volcano plot was built from the −log10 transformed false discovery rate (FDR) adjusted p-values and the log2 fold change values for each gene using the R package ggplot2 version 3.5.1; the significance cutoff was set to a −log10 transformed FDR adjusted p-value of 15 and a log2 fold chance below −1.5 or above 1.5.

#### Determination of myeloid cell origin

Two genes’ signatures were built from a set of 3 genes specific to either chromosome X (*XIST*, *JPX* and *FTX*, thereafter designated as “female signature”) or chromosome Y (*DDX3Y*, *KDM5D* and *USP9Y*, thereafter designated as “male signature”) and a module score was calculated on biopsy-derived scRNASeq samples for each signature using the R package UCell version 2.6.2*^30^*. Datasets showing a sex mismatch between the donor and the recipient with a clear opposing signatures expression observed using the blend parameter of Seurat’s FeaturePlot function were selected for further analysis: TAC1, TAC2, TAC3, TAC4, BELA2, BELA3, GSM4339776, GSM4339779, EXT241, NEPH012, NEPH016, NEPH017 and NEPH019. Cell identities were obtained from the blended FeaturePlot’s ggplot2 object using ggplot2’s ggplot_build function and then matching the data (x and y coordinate values of the scatterplot) of colored cells positive for either signature with UMAP’s cell embeddings.

### Cell-cell communication analysis

The R package CellChat version 2.1.2*^31^* was used to investigate ligands-receptors interactions between cell types and unravel communication networks. Annotations from reintegrated myeloid cells were transferred onto the same cells in the main object (matching cell barcode IDs from both objects) using Seurat’s SetIdent function. The CellChat database was appended to add interactions or modify existing interactions based on published data*^32–34^*. A CellChat object was created from the normalized count matrix and annotations of the main Seurat object and overexpressed genes, interactions, and communication probabilities were computed using CellChat’s respective functions identifyOverExpressedGenes, identifyOverExpressedInteractions, computeCommunProb (with TriMean type), filterCommunication (minimum 10 cells), computeCommunProbPathway, aggregateNet and netAnalysis_computeCentrality. Incoming and outgoing interaction strength were visualized using CellChat’s netAnalysis_signalingRole_scatter function. Networks of ligands-receptors pairs were generated using the cc_network function from the R package CCPlotR version 0.99.3*^35^*. Two other CellChat objects were created by splitting the normalized count matrix into two matrices, based on the rejection status (no rejection vs rejection). Overexpressed genes, interactions, and communication probabilities were computed again using the same functions from CellChat, and both objects were then merged together. A word cloud representing the most overexpressed ligands in each condition was built using CellChat’s computeEnrichmentScore function.

### Trajectory inference analysis

The R package slingshot version 2.10.0*^36^* was used to explore trajectories/lineages in myeloid cells. The cell embeddings matrix corresponding to UMAP coordinates of reintegrated myeloid cells was used as input for pseudotime inference using slingshot’s slingshot function, with classical monocytes as the starting cluster. Custom functions were developed for trajectories visualization on UMAP coordinates using ggplot2, the R package patchwork version 1.2.0 and slingshot’s slingPseudotime and slingCurves functions to extract inference results data for plotting.

### Trajectory-based differential expression analysis

The R package tradeSeq version 1.16.0*^37^* was used to find key genes significantly associated with each trajectory identified by slingshot. The raw count matrix of reintegrated myeloid cells was filtered using the RightOmicsTools’ tradeSeqPreprocess function, keeping genes only expressed in more than 10 cells and removing mitochondrial, ribosomal and non-coding genes, in order to reduce computational burden without losing much information, poorly represented genes being unlikely to contribute; 13,981 genes passed the selection. The filtered raw counts matrix as well as the slingshot output converted into a SlingshotDataSet object were first used as input to tradeSeq’s evaluateK function, which determines the number of knots to fit on each trajectory for differential expression analysis. Upon selecting 5 knots based on evaluteK results, the same input objects were then used to fit a negative binomial generalized additive model (NB-GAM) for each gene on each trajectory using tradeSeq’s fitGAM function on the supercomputer facilities of the Mésocentre de Franche-Comté. Computation time was greatly reduced using parallel computing from BiocParallel version 1.36.0 and batchtools version 0.9.17 R packages; a custom Slurm template was created and used as input to BiocParallel’s BatchtoolsParam function to take advantage of the scheduler capacity to automatically divide calculations into individual jobs, submit them in parallel to cluster nodes, and retrieve results from within the R environment. Within lineage DEG comparison was performed using RightOmicsTools’ tradeSeqTests function, which is a wrapper around tradeSeq’s earlyDETest adding FDR adjusted p-value and tidying the data, between knot 3 and 4 with fitGAM results as input. A rank score was built from the results of each of these statistical tests, corresponding to (*Wald statistic rank*)*^2^* + (*mean or median log2 fold change rank*)² (as computed by the R base rank function, with ties randomly broken) for each gene on each lineage or within lineage comparison, and plotted alongside the log10 transformed Wald statistic values and log10 transformed mean or median log2 fold change values for each gene using a custom ggplot2 code. Genes corresponding to the 90th percentile and up in terms of rank score (as computed by the R package stats’s quantile function, with 0.9 probability) from each lineage or within lineage comparison were selected and visualized on Venn diagrams, produced using the R package venn version 1.12. Knots on trajectories were visualized on UMAP coordinates using tradeSeq’s plotGeneCount function and a custom ggplot2 code. Scatterplots, smoothed gene expression curves and pseudotime heatmaps were generated respectively by the RightOmicsTools’ curveSmoothers and heatmapSmoothers functions.

### Interrogation of the PROMAD atlas

The online atlas tool was used to retrieve relative expression data for *FCN1*, *LILRA2*, *LILRA5*, *LILRB2*, *LST1* and *LY6E* in kidney biopsies from the control or acute rejection groups (https://shiny.maths.usyd.edu.au/PROMAD/). Only the 21 rejection datasets including the 6 genes of interest were considered here. After selecting the datasets on the tool, the object in RDS format was downloaded, and the expression data corresponding only to the 6 genes of interest were extracted in R. These data were then statistically compared between groups using Prism software for each of the datasets indicated using two-tailed Unpaired t test.

### Determination of transcription factor activity

The R package decoupleR version 2.8.0*^38^* was used to infer the transcription factor (TF) activity between *SELENOP*+ macrophages and *CXCL10*+ macrophages. The reference database, called CollecTRI*^39^*, containing a curated collection of TFs and their transcriptional targets, was built using decoupleR’s get_collectri function. Cells corresponding to *SELENOP*+ macrophages and *CXCL10*+ macrophages were subsetted from the reintegrated myeloid cells and the normalized count matrix as well as the CollecTRI object were used as input to the decoupleR’s run_ulm function, which computes a score using a Univariate Linear Model (ULM) for each TF in each cell based on the expression level of each TF’s transcriptional targets. This score reflects therefore the activity of a transcription factor based on the expression of the genes it regulates. The ULM score results of 1318 TFs were stored as a matrix in a new assay in the macrophages’ Seurat object and scaled and centered using Seurat’s ScaleData function. The top 10 most differentially active TF in each of the two macrophage’s subtypes were determined by averaging each TF activity in each cell type, subtracting the mean *SELENOP*+ macrophages activity from the mean *CXCL10*+ macrophages activity and selecting the first 10 and the last 10 TFs. Finally, the RightOmicsTools’ Cell_Heatmap function was used to visualize in the ULM assay the activity of these TFs in each cell.

### Deconvolution

The cellular composition of 1,374 publicly available kidney allograft biopsy microarray samples were inferred by the CIBERSORTx algorithm*^40^* using the kidney allograft biopsy scRNAseq data from this work.

#### scRNAseq signature matrix construction and validation

Annotations from reintegrated myeloid cells identified as classical monocytes, intermediate and non-classical monocytes, as well as *SELENOP*+ macrophages and *CXCL10*+ macrophages were transferred onto the same cells in the main object (matching cell barcode IDs from both objects) using Seurat’s SetIdent function. Cells originating from kidney allograft biopsies were subsetted from the main object and the raw count matrix was normalized to Counts Per Million (CPM) with Seurat’s NormalizeData function using the Relative Counts (RC) normalization method and a scale factor of 1,000,000. The benchmarking_init function from the R package deconvBenchmarking version 0.1.0*^41^* was used to divide the normalized count matrix into a training matrix, composed of 5% of cells randomly selected from each cell type, thus maintaining the original proportions of cell types within the original matrix, and a validation matrix, created using the remaining 95% of cells, whose expression was averaged per cell type and randomly distributed into 100 simulated pseudobulk mixtures (constituting the ground truth). The random cell type composition is known and varies for each of the 100 mixtures to benchmark CIBERSORTx performance. The training and validation matrices were exported as tab-separated text files using respectively the create_cibersortx_input parameter of deconvBenchmarking’s benchmarking_init function and RightOmicsTools’ Mixture_File_Builder function, and uploaded into the web interface of CIBERSORTx. The training matrix was used as input to the “Create Signature Matrix” module of CIBERSORTx. Given that a droplet-based technique (10X Chromium) was used for generating the scRNAseq datasets, the *Min. Expression value* parameter was reduced to 0.25. This adjustment aims to increase the reliability of the signature matrix, as recommended by CIBERSORTx, resulting in a matrix consisting of 6172 genes and 23 cell types. The signature matrix and the validation matrix were then used as input to the “Inpute Cell Fractions” module of CIBERSORTx to estimate the cell proportions of the simulated pseudobulk mixtures, using *S-mode* batch correction and 100 permutations for statistical significance. A Pearson’s correlation coefficient was calculated for each cell type by comparing the estimated cellular composition from CIBERSORTx to the simulated pseudobulk mixtures’ ground truth in order to validate the signature matrix.

#### Cellular composition estimation of microarray data

Six publicly available microarray datasets were downloaded from the NCBI’s GEO FTP servers (accession numbers: GSE21374*^42^*, GSE36059*^43^*, GSE48581*^44^*, GSE98320*^45^*, GSE147089*^46^* and GSE147451*^47^*) as gunzipped text files. After extraction, the text files were divided into metadata and expression matrix. For the GSE36059 and GSE48581 datasets, samples corresponding to ABMR, TCMR and Mixed were selected. For the GSE98320 dataset, the Borderline and ABMRsusp. samples were excluded. For the GSE147089 dataset, two samples with missing survival data were excluded. The probe ID of each gene was converted into its matching ENSEMBL ID using the R package oligo version 1.66.0 and the corresponding Affymetrix database. The six expression matrices were then loaded into BIOMEX software version 1.0.5*^48^* to map the ENSEMBL IDs into HGNC symbols. Finally, since the expression matrices were already log2-RMA normalized, they were not further processed and were exported and converted into tab-separated text files using the RightOmicsTools’ Mixture_File_Builder function and uploaded into the web interface of CIBERSORTx. The validated signature matrix and each expression matrix were then used as input to the “Inpute Cell Fractions” module of CIBERSORTx, using *S-mode* batch correction and 100 permutations for statistical significance. Various custom ggplot2 codes were employed to visualize the cell proportions as box plots, bar plots, scatter plots and violin plots. Radar plots were generated using the radarchart function from the R package fmsb version 0.7.6.

### M1 and M2 macrophages bulk RNA-sequencing data analysis

Raw data were downloaded from the NCBI’s GEO database (accession number: GSE146028*^49^*) and loaded into BIOMEX software version 1.0.5*^48^*. Metadata corresponding to macrophages M1 and M2 subtypes were added, counts were normalized. A volcano plot of M1 vs M2 comparison was generated, DEG results used to build the plot (log2 fold change, p-value and FDR adjusted p-value) were exported into R and the volcano plot was reconstructed from these values using ggplot2; the significance cutoff was set to a −log10 transformed FDR adjusted p-value of 2 and a log2 fold chance below −1.5 or above 1.5. Genes found to be significant on the volcano plot between *SELENOP*+ and *CXCL10*+ macrophages were represented and four quadrans were drawn depending on significance cutoff, with a fifth area representing no significance.

### Isolation of PBMC by density gradient

Peripheral blood from Buffy coats (Etablissement Français du Sang-BFC, France) was filtered through a 70µm strainer. Two volumes of blood were poured onto one volume of Lymphocyte separation medium (Eurobio Scientific, Les Ulis, France). Cells were then centrifuged for 30 min at 800g at room temperature. PBMC were washed with Phosphate-Buffered Saline (PBS, Thermofisher, Courtaboeuf, France) by centrifugation at 450g at 4°C for 5min. The cell pellet was resuspended in PBS, then centrifuged at 2100g for 2min at 4°C to remove the platelets.

### Flow cytometry analysis

Samples to be analyzed by flow cytometry were centrifuged at 700g 5min at room temperature. One million cells from each condition were labeled with fluorochrome-coupled antibody for 15min at 4°C. Prior to cytometric analysis, labeled cells were washed with 2mL PBS and centrifuged at 700g 5min at room temperature before being resuspended in 200 µL PBS for analysis on a DxFlex cytometer (Beckman Coulter, Paris, France) or an Attune cytometer (Thermofisher). The following antibodies were used: anti-LILRB2 PE-Vio 770 (REA184, Miltenyi Biotec), anti-CD16 BV605 (3G8, Biolegend). When indicated, incubation with Fixable Viability Stain 700 (BD Biosciences, Paris, France) was performed prior to incubation with the coupled antibodies according to the supplier’s recommendations.

### Monocyte sorting

Monocytes were magnetically sorted from PBMC either by the CD14 Microbeads kit for transduction experiments or by the PanMonocyte kit (Miltenyi Biotec, Paris, France) for in vitro activation experiments following the supplier’s recommendations. A minimum purity of over 85% was required to start the experiments.

### Monocyte activation

Anti-CD47 (PA5-80435, Thermofisher) and a non-specific control antibody (10500C, Thermofisher) were coated in a 12-well plate (3413, Corning) for 4h at 37°C at a concentration of 3µg/mL in PBS. The wells were then emptied and 0.5.106 purified monocytes suspended in 1mL of warm RPMI supplemented with 10% FBS and 1% Pen/Strep medium were deposited in each well before spinoculation for 1min at 100g, followed by incubation at 37°C with 5% CO2.

### Lentiviral transduction

To induce expression of the LILRB2 in THP-1, we used lentiviral strategy (VectorBuilder, Neu-Isenburg, Germany) with a lentivirus encoding *LILRB2* and a reporter gene encoding *TagGFP2*. THP-1 were transduced at a multiplicity of infection (MOI) of 2.5 in the presence of 8 mg/L Polybrene (PL001, VectorBuilder). Briefly, 0.5.10^6^ cells per well were seeded in P24 well plate in the presence of virus particles and Polybrene. Control wells contained only 0.5.10^6^ cells/well and culture medium. The transduction was performed by spinoculation, i.e. centrifugation at 32°C, 1000 g for 90 minutes before culture for 2 days. To induce expression of the LILRB2 in human primary macrophages, monocytes freshly isolated from healthy volunteers were subjected to M0 differentiation by M-CSF (130-096-492, Miltenyi Biotec) at 25ng/mL in X-Vivo medium (BEBP02-054Q, Lonza, Colmar, France) supplemented with 10% of FBS (10271-106, Gibco) for 2 days before transduction with virus-like particles packaging Vpx to overcome virus restriction*^50^*. The production of Vpx particles was performed as previously described*^51^*. At day 4 of culture, the cells were transduced with the lentivirus encoding *LILRB2* and *TagBFP2* at MOI 1. At day 10 of culture, the fully differentiated macrophages were collected for cytometry analysis and LILRB2-HLA interactions assay.

### FACS isolation of THP-1 clones

Cell sorting of transduced THP-1s was performed using a BDFACS ARIA III sorter (BD Biosciences, Paris, France) at the ImaFlow Platform (Dijon, France). THP-1 cells were labelled with an anti-LILRB2 PE Vio 770. The clones expressing different levels of the reporter gene TagBFP2 were selected and sorted in a 96-well plate pre-filled with culture medium to obtain one single clone per well. Identification of each clone according to the expression levels of LILRB2 was performed on FlowJo software using the Index Sort plugin (BD Biosciences).

### LILRB2-HLA interaction assay

In order to assess LILRB2-HLA interaction, cells of interest were incubated with HLA-B27-APC pentamer (F294-4B-E, ProImmune, Oxford, UK) or HLA-A2-A2-PE pentamer (F10-2B-E, ProImmune) according to the supplier’s recommendations.

### ORLY-EST ancillary cohort

ORLY-Est (Orientation of the Lymphocyte Response to the Occurrence of Atherosclerotic Complications After Kidney Transplantation, NCT02843867) is a prospective cohort study of kidney transplant patients included in seven French transplant centers (Besançon, Clermont-Ferrand, Dijon, Le Kremlin-Bicêtre, Nancy, Reims and Strasbourg), whose main objective was to understand the interactions between immune status and post-transplant outcomes. For each patient, blood samples were collected on the day of transplantation (d0) and one year later (d365). Peripheral blood mononuclear cells (PBMC) were separated by density gradient centrifugation (Pancoll, Pan-Biotech GmBH Aidenbach, Germany) and stored in liquid nitrogen at the Centre de Ressources Biologiques Ferdinand Cabanne (Dijon, France). Samples were collected with the regulatory approval of the French Ministry of Health (agreement number DC-2008-713 dated June 11, 2009), and the study was approved by the ethics committee of the Université de Franche-Comté in 2008. All patients included in the ORLY-Est study were of legal age at the time of transplantation, and gave written informed consent. For this ancillary present study, PBMC were stained for flow cytometry analysis as previously described in an 8-batch sequence including a common healthy PBMC for batch normalization. To ensure analysis quality, cytometry files were normalized between batches using the CytoNorm*^52^* plugin, and abnormal events intrinsic to cytometry were removed using the FlowClean*^53^* plugin.

### Inflammation level analysis

A diagnostic label was awarded to each biopsy based on the presence and severity of these histological lesions and on the DSA status. In this aim, all biopsies were categorized into one of six recently described clinicopathological categories*^54^*, based on the presence of acute histological lesions and recipient DSA status (available at https://rejectionclass.eu.pythonanywhere.com/). This data-driven classification allows for visualization of the biopsies on a two-dimensional polar plot in which the theta angle associates with the spatial localization of the inflammation, i.e., microvascular inflammation versus tubulointerstitial inflammation. The radius indicates the global severity of the inflammatory infiltrate given by the sum of re-weighted acute lesions scores, scaled to the unit interval (from 0 to 1) and is hereafter designated as “Activity index”*^54,55^*.

### Autologous T Cell Proliferation Assays

On the day of monocyte sorting, the remaining PBMC from the same donors were cultured in complete RPMI supplemented with IL-2. To achieve coculture with syngeneic macrophages, PBMCs were stained with Cell Trace Red according to the manufacturer’s procedure (Cell Trace, Thermofisher). Labeled cells were cultured with human CD3/CD28 T activator beads (Dynabeads, Thermofisher) in a 96-well UBottom plate in 100µL of complete RPMI at 1,10^6^ cells/mL with or without 100µL of macrophages. For dose-dependent experiments, the number of labeled PBMC was fixed, while the number of macrophages varied from a ratio of 1:1 to 1:4 (macrophages:PBMC). T-cell division was detected after 5 days by flow cytometry and analyzed using the Proliferation module of FlowJo software.

### Statistical analysis

We report descriptive statistics using mean and standard deviation (or median and interquartile range for skewed distributions) for continuous variables or numbers, and percentages for discrete variables, for the full cohort and for the rejection subgroups. We used the most recent (as of July 2025) versions of all software programs, including R Studio (version 2025.05.0+496) and Prism (version 10; GraphPad Software, San Diego, CA, United States) for statistical analysis and data presentation. The time-dependent ROC curve analysis of survival data was performed with the function survivalROC from the R package survivalROC version 1.0.3.1*^56^* at 365 days post biopsy and using the Kaplan-Meier (KM) method. The Youden index*^57^* was computed from the specificity and sensitivity values of the model, as defined by the formula *sensitivity* + *specificity* - 1 for each cell type and represented using a custom gpplot2 code. The Kaplan-Meier survival curves*^58^* right-censored at 1825 days post biopsy were generated using the survfit and Surv functions from the R package survival version 3.5-7*^59^* and visualized with the ggsurvplot function from the R package survminer version 0.5.0*^60^*. Cox proportional hazards analysis of survival data right-censored at 365 days post biopsy was performed using survival’s coxph function and visualized using survminer’s ggforest function.

## Results

### Single-cell integration of kidney allograft and blood-derived cells

To characterize the infiltration of circulating monocytes and their differentiation into macrophages in kidney transplant patients, we integrated 46 single-cell RNA-Seq datasets, including 13 datasets corresponding to blood-derived cells (including N=5 without rejection and N=8 rejections) and 33 datasets corresponding to kidney allograft-derived cells (including N=13 without rejection and N=20 rejections, Fig. 1A). The rejection phenotypes were diverse from a histological point of view and showed varying levels of inflammation, ranging from mild to severe. In addition, for N=3 patients, including two cases of rejection, we were able to integrate both blood cells and cells from the kidney allograft collected on the same day, contributing to the robustness of the results (Fig. 1B). After passing quality controls (Fig. S1a), we successfully integrated 150,876 transcriptomes without any major batch effects, whether between datasets, tissue of origin, or between the rejection group compared to the non-rejection group (Fig. S1b). In an unsupervised manner, 23 different clusters were identified, corresponding to blood and kidney cells. The annotation using canonical and unsupervised markers (Fig. 1C, Supplemental Table 1) of these clusters led to the identification of immune cells subtypes such as *S100A12*+ *LYZ*+ *CD14*+ monocytes and *C1QA*+ *C1QB*+ *MS4A4A*+ macrophages (Fig. 1D). When comparing the proportion of immune cells between the rejection and non-rejection groups within the allograft, an increase in the proportion of immune cells was observed in cases of rejection, as expected. Notably, the proportion of macrophages within the allograft increased from 1.19% in non-rejection to 3.95% in cases of rejection (Fig. 1E). A global, unsupervised comparison of the main cellular interactions between rejection and non-rejection on all integrated cells reveals that the no rejection group is mainly associated with CCL5 and LGALS9 (Fig. 1F). In contrast, the rejection group is predominantly associated with CXCL10 and CXCL9 in line with recently reported pan-organ transcriptomic data across organs, which confirms that CXCL9 and CXL10 chemokines are central in rejection process*^3^* and underscore the ubiquitous role of myeloid cells in solid-organ transplantation*^4^*.

**Fig. 1.**
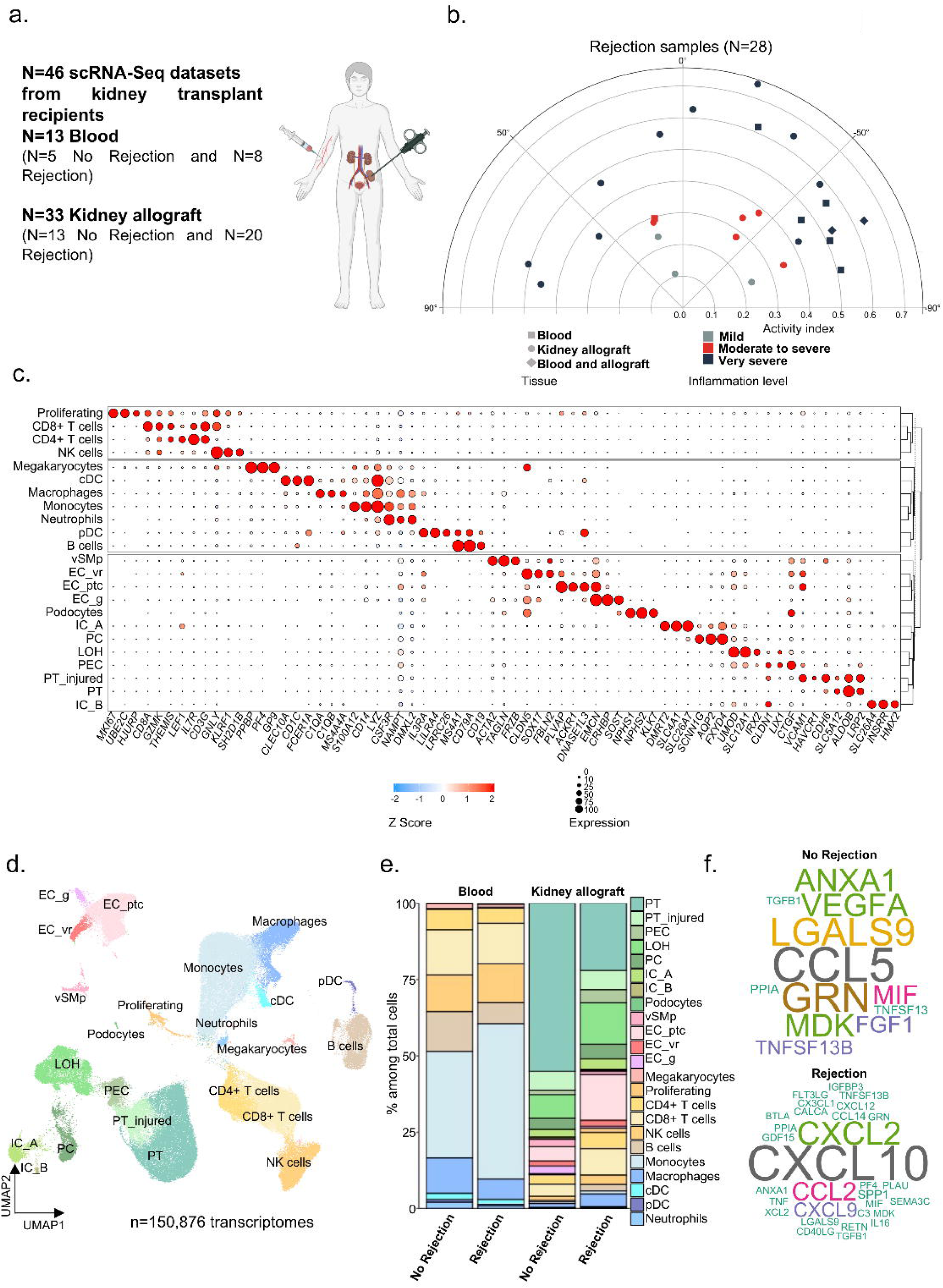
Single-cell integration of kidney allograft and blood-derived cells. a. Experimental Approach. 46 scRNAseq datasets from kidney transplant patients were used for this analysis (n□=□13 blood with 5 no rejection and 8 rejection, and n = 33 kidney allograft biopsies with 13 no rejection and 20 rejection). b. Polar plot of the 28 rejection samples showing tissue origin and estimated inflammation level severity. c. Clustered dot plot showing average gene expression values of canonical markers (log normalized) and expression percentage of cell types represented on the Uniform Manifold Approximation and Projection (UMAP) plot. d. UMAP plot of 150,876 cells passing QC filtering and doublets removal. The main kidney cell types are represented, including loop of Henle (LOH), podocytes, vascular smooth muscle and pericytes (vSMp), proximal tubule (PT), intercalated cells type A and B (IC_A / IC_B), three endothelial cell subsets comprising vasa recta (ECvr), glomerular (ECg) and peritubular capillaries (ECptc), myeloid cells, lymphoid cells, megakaryocytes, and proliferating cells. e. Stacked barplots representing the relative cell proportion of each cell type on the UMAP plot, split by tissue origin (blood or kidney allograft biopsies) and by outcome (no rejection or rejection). f. Word cloud obtained using the computeEnrichmentScore function of Seurat and split by outcome (no rejection or rejection).

### A pro-inflammatory macrophage subpopulation expressing *CXCL10* is specifically enriched during rejection

We then selected all the myeloid cells annotated previously as ‘Monocyte’, ‘Macrophage’ and ‘cDC’ (Fig. 1D) for reintegration, in order to better understand the heterogeneity of myeloid cells, both in the blood and infiltrating the allograft after kidney transplantation. We thus obtained 11 different clusters in an unsupervised manner (Fig. 2A). Among them, we identified in an unsupervised approach 6 monocyte populations and their specific markers (Supplemental Table 2), such as *S100A12*+ *CD14*+ *LYZ*+ classical monocytes, a *MARCO*+ *APOBEC3A*+ population of intermediate monocytes, an *IFIT1*+ *HERC5*+ *MX1*+ population of IFN-activated monocytes, a *FCGR3A*+ *CDKN1C*+ non-classical monocyte population, a *PECAM1*+ *CX3CR1*+ *TCF7L2*+ patrolling monocyte population, and a monocyte population differentiating into dendritic cells, the mo-DCs expressing *AHNAK* and *HIST1H1E*. In addition, 3 populations of dendritic cells were detected: a *CLEC10A*+ *CD1C*+ *FCER1A*+ conventional type 2 DC (cDC2), a *CLEC9A*+ *XCR1*+ *GCSAM*+ cDC1 population and a *TNF*+ *CCL3L1*+ *CD83*+ population named hereafter “CD83 mature DC”. Finally, 2 subtypes of macrophages were distinguished from all other myeloid cells: a *SELENOP*+ *STAB1*+ *TREM2*+ population and a *CXCL10*+ *ANKRD22*+ population (Fig. 2B). Concerning the proportion of the overall myeloid compartment detected within allografts, we observed a substantial increase with a doubling of the global myeloid population during rejection (Fig. 2C) in line with previous reports*^2,61^*. More specifically, the infiltration ratio of myeloid cells differed according to their subtypes. While the proportion of cDC1, cDC2, CD83+ DC, mo-DC and patrolling monocyte was not enriched in allograft, the proportion of *CXCL10*+ macrophages, classical, intermediate and IFN-activated monocytes and *SELENOP*+ macrophages was statistically increased in rejection. In particular, a 5-fold increase in the proportion of *CXCL10*+ macrophages was observed, indicating a major enrichment of this population within the kidney allograft during rejection (Fig. 2D). Among myeloid cells detected solely within the allograft, the proportion of *CXCL10*+ macrophages increased significantly in rejection (0.0% vs 6.4%, P=0.0009) compared with *SELENOP*+ macrophages (29.2% vs 25.4%, P=0.68) suggesting that not only infiltration but also differentiation of this population is directly associated with the allogeneic response against the graft (Fig. 2E).

**Fig. 2.**
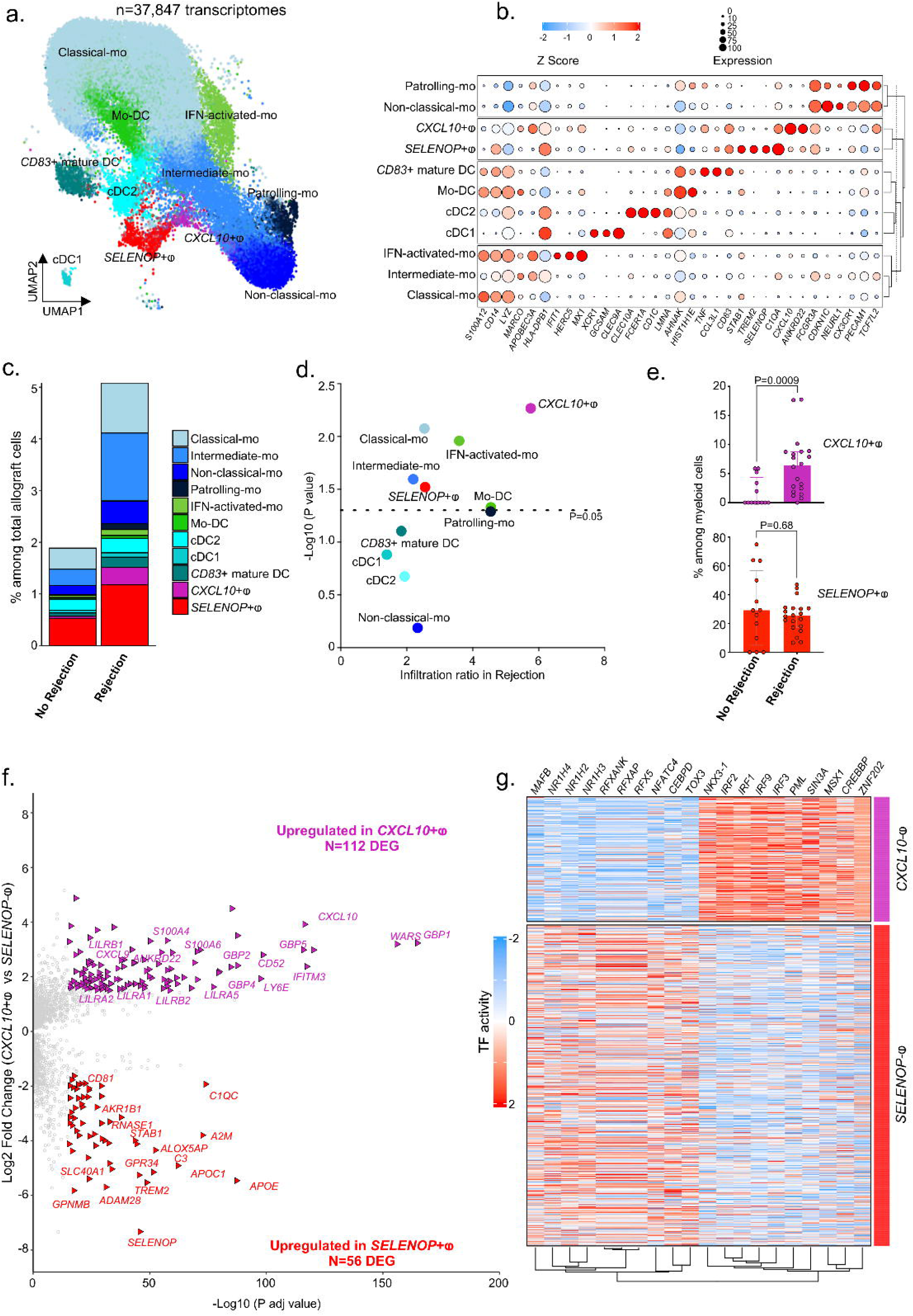
A proinflammatory macrophage subpopulation expressing *CXCL10* is specifically enriched during rejection. a. UMAP plot of 37,847 cells following Seurat’s reintegration of monocytes, macrophages and cDC cell types from the main object. The main myeloid cell types are represented, including the three expected subsets of monocytes (Classical-mo, Intermediate-mo and Non-classical-mo) as well as more specific subtypes such as classical monocytes activated by interferon-gamma (IFN-activated-mo), classical monocytes in the process of differentiating into monocyte-derived dendritic cells (Mo-DC), and non-classical monocytes expressing higher levels of adhesion molecules (Patrolling-mo). Three subtypes of dendritic cells were also found, with both expected main conventional subsets (cDC1 and cDC2) and a *CD14*+ *CD83*+ cluster of monocyte-derived mature dendritic cells (*CD83*+ mature DC). Finally, two macrophages’ subtypes were identified, one expressing high levels of *SELENOP* (*SELENOP*+L) and another expressing high levels of several chemokines (*CXCL10*+L). b. Clustered dot plot showing average gene expression values of canonical markers (log normalized) and expression percentage of major cell types represented on the UMAP plot. c. Stacked barplots representing the relative cell proportion of each myeloid cell type among all kidney allograft cells, split by outcome (no rejection or rejection). d. Scatterplot representing the relative proportion ratio of each myeloid cell type among all kidney allograft cells in no rejection versus rejection samples, defining an infiltration score for each myeloid cell type during rejection. e. Barplots showing the relative proportion of *SELENOP*+ and *CXCL10*+ macrophages among all myeloid cells in no rejection versus rejection samples. Two-tailed Mann–Whitney tests comparing No rejection and Rejection samples indicate significant difference in *CXCL10*+ macrophages proportions but not in *SELENOP*+ macrophages proportions. Two-tailed Mann Whitney tests were performed to compare the groups. P-values are shown. f. Volcano plot representing DEG between *SELENOP*+ and *CXCL10*+ macrophages. g. Heatmap representing the transcription factor activity regulating the DEG between *SELENOP*+ and *CXCL10*+ macrophages.

**Fig. 3.**
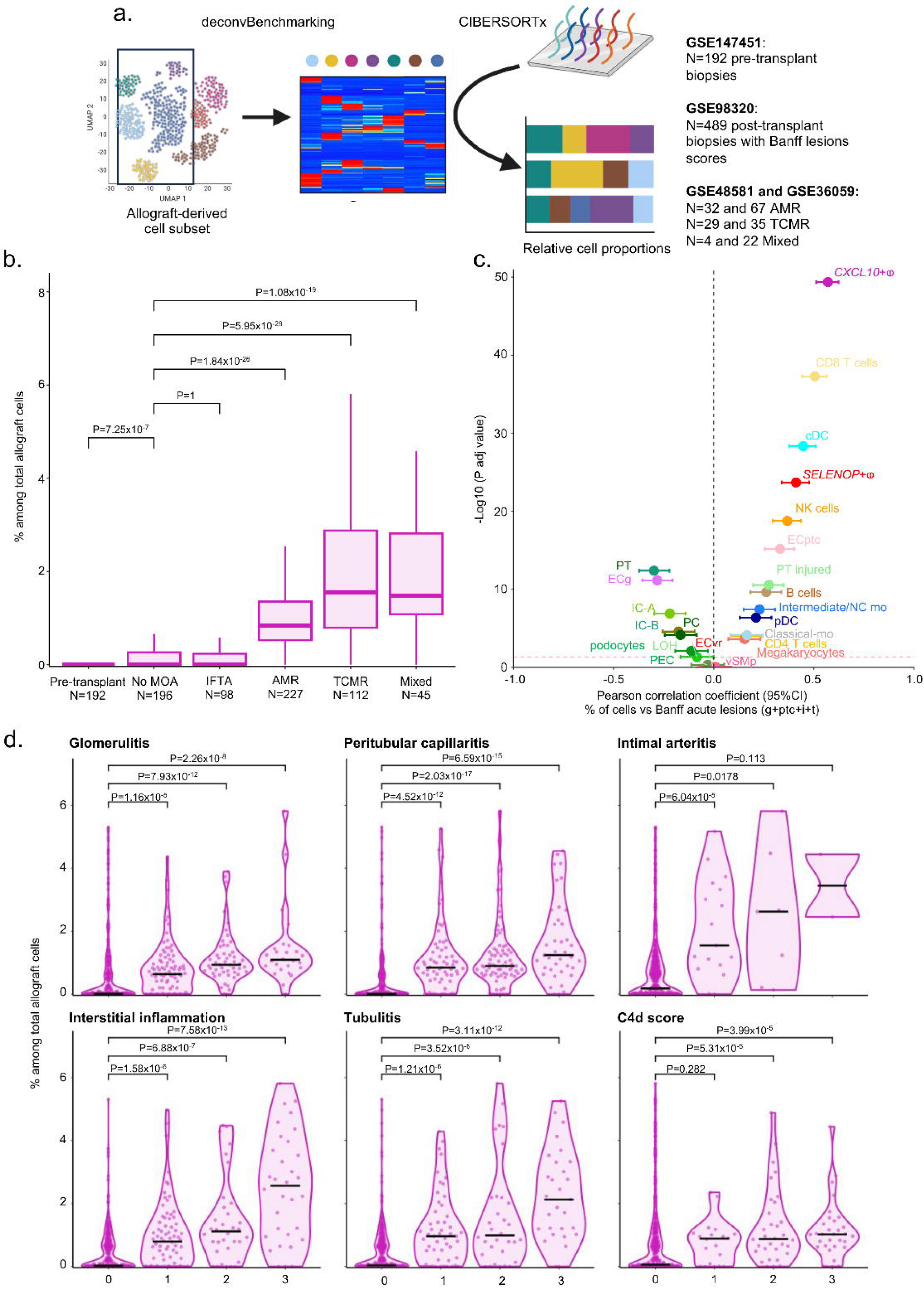
*CXCL10*+ macrophages are absent pre-transplantation and dramatically increase upon acute rejection. a. Experimental approach. scRNAseq-derived signature matrix was used for deconvolution of the dataset GSE147451 encompassing 192 transcriptomes from pre-transplant kidney biopsies, the dataset GSE98320 encompassing 489 transcriptomes from post-transplant biopsies with rejection phenotypes and Banff lesions scores as well as the datasets GSE48581 and GSE36059 encompassing 189 transcriptomes from post-transplant biopsies with rejection phenotypes. b. Frequency of *CXCL10*+ macrophages within the allograft inferred by deconvolution in GSE147451, GSE98320, GSE48581 and GSE36059 and stratified according to clinical outcome. No MOA = No Major Abnormality, IFTA = Interstitial Fibrosis and Tubular Atrophy, AMR = Antibody-Mediated Rejection, TCMR = T Cell-Mediated Rejection. The difference between groups was assessed by a two-tailed Kruskal-Wallis test and multiple comparisons using the Dunn’s test. P-values are shown. c. Pearson’s correlation coefficient and −log10 of the associated p-value, represented by a dot, with the 95% confidence interval indicated by error bars, for the correlation between the sum of Banff lesion scores related to acute rejection and the frequency of different immune cells within the allograft inferred by deconvolution in GSE98320. d. Frequency of *CXCL10*+ macrophages within the allograft inferred by deconvolution in GSE98320 and stratified according to Banff lesion scores of acute rejection. The difference between groups was assessed by a two-tailed Kruskal-Wallis test and multiple comparisons using the Dunn’s test. P-values are shown.

**Fig. 4.**
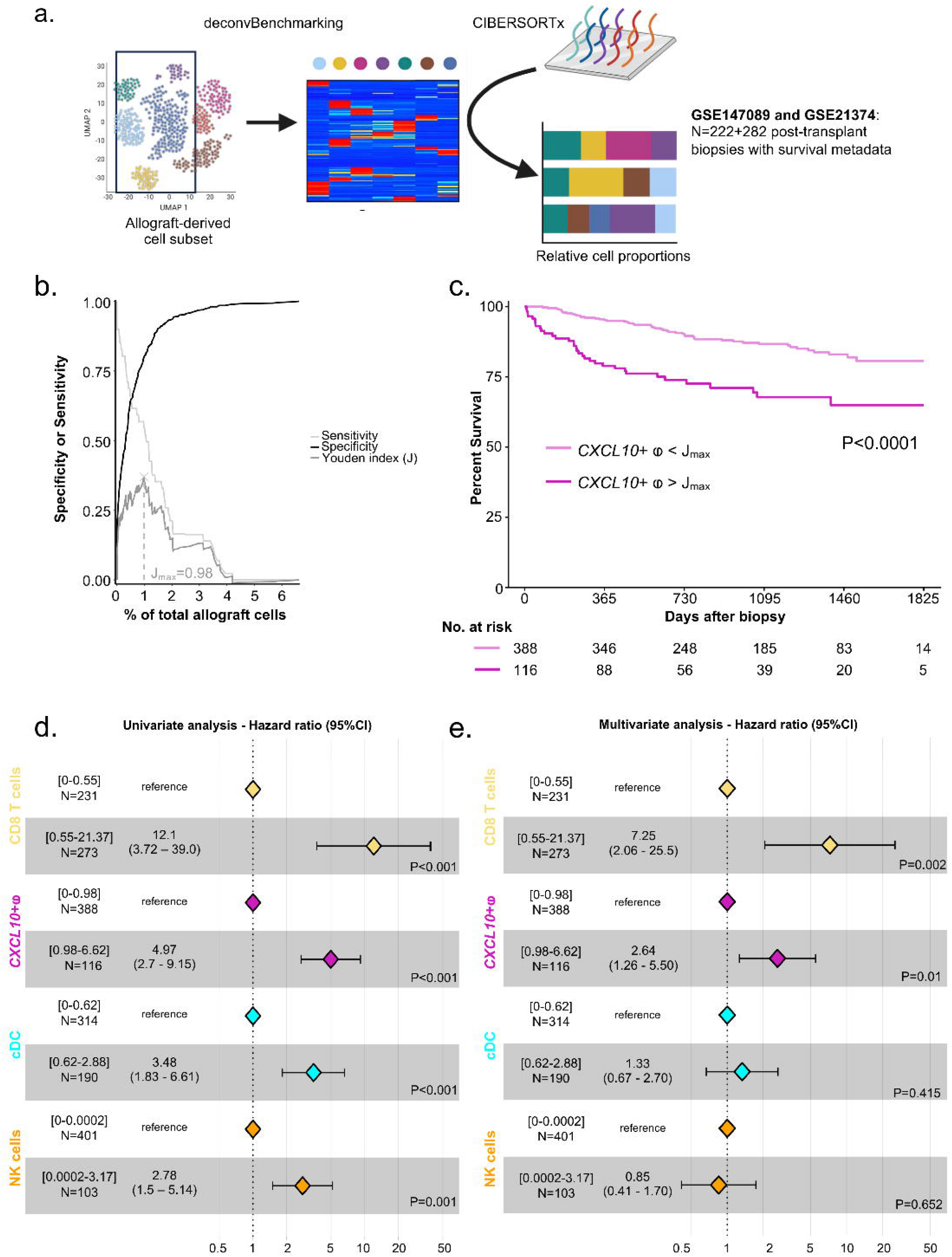
*CXCL10*+ macrophages infiltration is associated with graft loss, independent of other immune cells. a. Experimental approach. scRNAseq-derived signature matrix was used for deconvolution of the datasets GSE147089 and GSE21374 encompassing 504 transcriptomes from post-transplant biopsies with survival metadata. b. After evaluating the diagnostic accuracy of *CXCL10*+ macrophages infiltration for predicting graft loss using time-dependent ROC curve analysis at one-year post biopsy, the optimal cutoff of the *CXCL10*+ macrophages proportion that maximizes the Youden index was calculated from the sensitivity and specificity values of the model (Jmax=0.984%). c. Kaplan Meier curves of allograft survival data right-censored at five-years post biopsy based on *CXCL10*+ macrophages proportion, stratified in two groups using the calculated Youden index cutoff at one-year post biopsy. The difference in survival between the two groups was assessed using the log-rank test. P-value is shown. d-e Univariate and multivariate Cox proportional hazards analysis of allograft survival data right-censored at one-year post biopsy, with the proportion of the four cell types most-associated with the activity index inflammation within the allograft as predictors, stratified in two groups using their respective calculated Youden index cutoffs at one-year post biopsy. P-values are shown.

### *CXCL10*+ and *SELENOP*+ macrophages harbor distinct transcriptomic programs

By comparing the transcriptomic profiles of the two macrophage populations (*CXCL10*+ and *SELENOP*+), we observed that *CXCL10*+ macrophages also overexpressed 112 signature genes, including proinflammatory genes such as *WARS*, *CXCL9*, *GBP1*, *GBP2* and *GBP5* which have previously been described as major biopsy-derived transcripts associated with rejection*^62^* (Fig. 2F). It is noteworthy that among these signature genes of *CXCL10*+ macrophages, a third is included in the Banff Human Organ Transplant (B-HOT) probe set panel*^63^* including pertinent genes related to rejection and innate and adaptive immune responses (N=36/112, Supplemental Table 3). Moreover, they specifically express genes encoding LILR receptors, some of which, such as LILRB1 and LILRB2, can bind HLA molecules*^64^*. We then identified the transcription factors (TFs) potentially involved in DEG regulation in both sub-populations. In particular, *SELENOP*+ macrophages express TFs from the NR1H family of nuclear receptors, namely *NR1H4* (FXR), *NR1H3* (LXRα) and *NR1H2* (LXRβ), which regulate lipid homeostasis*^65^*. In turn, *CXCL10*+ macrophages strongly express TFs of the Interferon Regulatory Factors (IRFs) family, notably *IRF3*, which induces *CXCL10* expression independently of IFN-γ activation*^66^*. In a reductive manner, macrophage subtypes have historically been stratified according to pro-inflammatory M1*^67^* and pro-fibrotic M2*^68^* profiles obtained after in vitro differentiation from monocytes. We compared the transcriptomic profiles of *CXCL10*+ and *SELENOP*+ macrophages identified in vivo with M1 and M2 macrophages generated in vitro*^49^*. In *CXCL10*+ macrophages, 71.8% of their signature genes corresponded to genes upregulated in M1, revealing around 30% of “discordant” genes, including S100A4 and S100A6, responsible for cell mobility. As for *SELENOP*+ macrophages, only 32.7% of their signature genes were shared with M2, suggesting that the historical stratification between M1 and M2 derived from in vitro experiments is too limited and cannot be directly matched to macrophage subtypes described in vivo*^69^* (Fig. S2a). We then examined the origin of the two types of macrophages using the expression of X-linked genes or Y-linked genes as previously described in patients who had received a transplant from a sex other than their own*^70^*. Unambiguously, we observed that 100% of *CXCL10*+ macrophages originated from the recipient, suggesting that the presence of these cells entirely relies on infiltration by circulating monocytes. *SELENOP*+ macrophages, on the other hand, comprise around 18% donor-derived cells and 82% recipient-derived cells, so partly tissue-resident (Fig. S2b). A comparison of the markers of these two subsets revealed that donor-derived *SELENOP*+ are *SELENOPhi* and *TREM2*-, while recipient-derived *SELENOP*+ are *SELENOPlow* and *TREM2*+ (Fig. S2c). In term of cellular interactions predicted at transcriptomic level between *CXCL10*+ and *SELENOP*+ macrophages and their microenvironment in the allograft, we detected more than 35 significantly enriched incoming signaling and more than 40 outgoing signaling (Fig. S2d). Among incoming signaling, we notably detected cell-contact CD47-SIRPA and LILRA1-B1-B2-class I HLA signaling which have been reported as responsible of allogeneic myeloid response in the animal*^71,72^* but also PECAM1-CD38, the interaction which is required for leukocyte transendothelial migration suggesting that the *CXCL10*+ macrophages derive from recipient-derived monocytes. In contrast, PECAM1-CD38 interaction was not detected in *SELENOP*+ macrophages suggesting that this population is less susceptible to derive from monocyte infiltrating the allograft through proinflammatory chemokine gradient (Fig. S2b). In term of cytokine-receptors, *CXCL10*+ macrophages mainly receive TNF signaling through TNF-TNFRSF1 but also chemokines through CCL-CCR1. In return, *CXCL10*+ macrophages outgoing signaling is characterized by CXCL-receptors signaling and TNF signaling, suggesting both cis and trans-signaling through this pathway. Altogether these results suggest that the *CXCL10*+ macrophages derive exclusively from the recipient and infiltrate the allograft to induce severe inflammation.

### *CXCL10*+ macrophages are specifically generated during the allograft response and cause acute rejection

In order to generalize our initial results to much larger external cohorts, we used the tool of deconvolution to estimate the fraction of *CXCL10*+ macrophages in pre- or post-transplant renal grafts. To this end, we reanalyzed datasets corresponding to N=192 preimplantation biopsies, N=489 post-transplant renal biopsies for which Banff lesions were available and N=189 post-transplant renal biopsies for which the rejection subtypes were available (Fig.3a). Strikingly, no *CXCL10*+ macrophages were detected in pre-transplant biopsies (median = 0.000% of total allograft cells), while a very limited contingent of these cells was measured in biopsies showing no major abnormalities (No MOA) or atrophy-fibrosis (IFTA) (median = 0.003 % and 0.000% respectively, Fig.3b). In contrast, biopsies depicting allogeneic rejection showed a significant increase of *CXCL10*+ macrophages with respectively a median of 0.84 % of *CXCL10*+ macrophages in AMR, 1.55 % in TCMR and 1.47% in mixed rejection. Conversely, in pre-implantation biopsies, the proportion of other myeloid cell types, such as *SELENOP*+ macrophages or cDCs, is at the same level as in No MOA, IFTA or AMR biopsies, suggesting that these cells are all detectable in the graft at the time of transplantation (Fig. S3a). These results confirm that, unlike other myeloid cells, *CXCL10*+ macrophages originate from the recipient and infiltrate the graft only during acute rejection and not in other contexts such as fibrosis. To clarify the association between *CXCL10*+ macrophage infiltration and the intensity of inflammation during rejection, we measured the rejection activity index as previously described*^54^*. We first observe considerable heterogeneity in biopsy profiles (Fig.S3b) and that, irrespective of rejection subtypes, the proportion of *CXCL10*+ macrophages measured in biopsies was strongly correlated with the acute rejection level given by the sum of g+ptc+i+t Banff lesions (Fig.3c) and inflammation given by the rejection activity index (Fig.S3c). We then compared the correlation between inflammation and other cell types, and found that *CXCL10*+ macrophages were the immune cells most positively correlated with inflammation, followed by NK, cDC and CD8 T cells (Fig.S3d). By stratifying biopsies according to Banff lesion scores characterizing rejection, the proportion of *CXCL10*+ macrophages gradually increase in the presence of glomerulitis, peritubular capillaritis, interstitial inflammation, intimal arteritis, tubulitis and C4d deposition (Fig.3d). Comparing to other myeloid cell types, their fold change in biopsies showing presence of acute rejection lesions was considerable (Fig.S3e). We then investigated whether *CXCL10*+ macrophage infiltration was associated with graft outcome. *CXCL10+* macrophage levels were measured by deconvolution in 504 post-transplant biopsies (Fig.4a). At 1-year post-biopsy, the maximum Youden index (J_max_) was calculated, indicating that a proportion of 0.984% *CXCL10*+ macrophages discriminated between patients with significantly different graft survival (Fig.4b). Biopsies were then stratified into two groups: those in which more than 0.984% *CXCL10*+ macrophages were found, and those in which less than 0.984% of allograft cells were *CXCL10+* macrophages. The detection in the biopsy of more than 0.984% of *CXCL10*+ macrophages infiltrating the allograft resulted in a significant decrease of the graft survival (Fig.4c, Log-rank P-value <0.0001). We then performed a univariate Cox regression and observed that the proportion of *CXCL10+* macrophages measured in the biopsy was significantly related to the risk of subsequent graft loss (hazard ratio, 4.97; 95% CI, 2.7 to 9.15; P<0.0001). Of note, NK cells, CD8 T cells and cDC, the other cell types associated with inflammation (Fig.S3d), were also significantly associated with the risk of graft loss. Importantly, we built a multivariate Cox regression model (Fig.4d) that confirmed that patients with high *CXCL10*+ macrophages proportion infiltrating the allograft had a higher risk of graft loss than those with fewer *CXCL10*+ macrophages (hazard ratio, 2.64; 95% CI, 1.26 to 5.5; P=0.01) independently of NK cells, CD8 T cells and cDC. Overall, these results suggest that, after transplantation, pro-inflammatory *CXCL10+* macrophages from the recipient infiltrate the allograft and induce microvascular and interstitial lesions, contributing to graft loss, independently of other immune cells.

### Myeloid cells differentiation is driven by six main trajectories in kidney transplant recipients

In order to better characterize the origin of the *CXCL10*+ macrophages, we next built unsupervised pseudotime trajectories among myeloid cells. In the interests of transparency and reproducibility, our entire monocyte trajectory construction pipeline is publicly available. All of these trajectories originated from blood-derived classical monocytes and formed a branch at the level of intermediate monocytes, before being distributed in clusters of fully differentiated cells such as DCs and macrophages within the allograft, or as non-classical or patrolling monocytes which were detected in both blood and allograft (Fig. 5a). These data suggest that classical monocytes can differentiate into intermediate and then non-classical monocytes as is also the case in the event of systemic inflammation*^73^*. Surprisingly, one of the trajectories formed a loop passing through CD83 mature DC and back to classical monocytes. Moreover, the six trajectories begin to separate at the branching point annotated “3” corresponding to intermediate monocytes. We therefore wanted to investigate which transcripts were specific to segment 3-4 of the branched section corresponding to *CXCL10*+ macrophages. For this purpose, we compared the transcripts associated with segment 3-4 of each trajectory (Fig. 5b) considering only the most specific transcripts (Rank score>q90). We intersected the transcripts of each comparison and defined 22 transcripts which were highly associated to segment 3-4 of CXCL10+ macrophages (Fig. 5c). Among these 22 transcripts, 6 of them, *FCN1*, *LY6E*, *LILRA2*, *LILRB2*, *LST1* and *LILRA5* shared a similar profile and high expression (Fig. 5d). In order to confirm the increased expression of these 6 genes during rejection in external global transcriptomics datasets, we queried the PROMAD atlas*^3^* (Fig. S4a). In 21 external datasets encompassing the 6 genes of interest and representing over 2,500 biopsies, *FCN1* and *LST1* expression was found to be systematically significantly increased in the graft during rejection except in two datasets. *LILRA2* was significantly increased in 18/21 datasets, and *LILRB2* in 15/21 datasets, confirming the importance of these transcripts during rejection. *LILRA5* and *LY6E*, on the other hand, were only significantly increased in 7/21 datasets indicating that they may be more specific to a subtype of immune response (Fig. S4b). Plotting the expression of these genes along trajectories, we confirmed the specificity of these transcripts the *CXCL10*+ macrophages trajectory. Surprisingly, we also observed that *LILRB2*, *LILRA5* together with *LILRA2* transcripts were upregulated at the end of the CD83 mature DC trajectory back to classical monocytes (Fig. 5e). In mice, myeloid allorecognition is primed by CD47/SIRP-17 axis*^71^* and leads to the overexpression of paired immunoglobulin receptors (PIRs) at myeloid cell surface*^72^*. The human orthologs of PIRs are leukocyte immunoglobulin-like receptors (LILRs). Here, our data suggest that the trajectory of *CXCL10*+ macrophages relies on three LILRs, *LILRB2*, *LILRA2* and *LILRA5*, but also that recirculating monocytes may retain the imprint of their passage through the allograft. We therefore expanded our analysis to the other LILRs and observed that *LILRA1*, *LILRB1* and *LILRB3* also follow the same expression profiles: they are strongly increased in *CXCL10*+ macrophages and they are upregulated at the end of the trajectory returning to classical macrophages (Fig. S5).

**Fig. 5.**
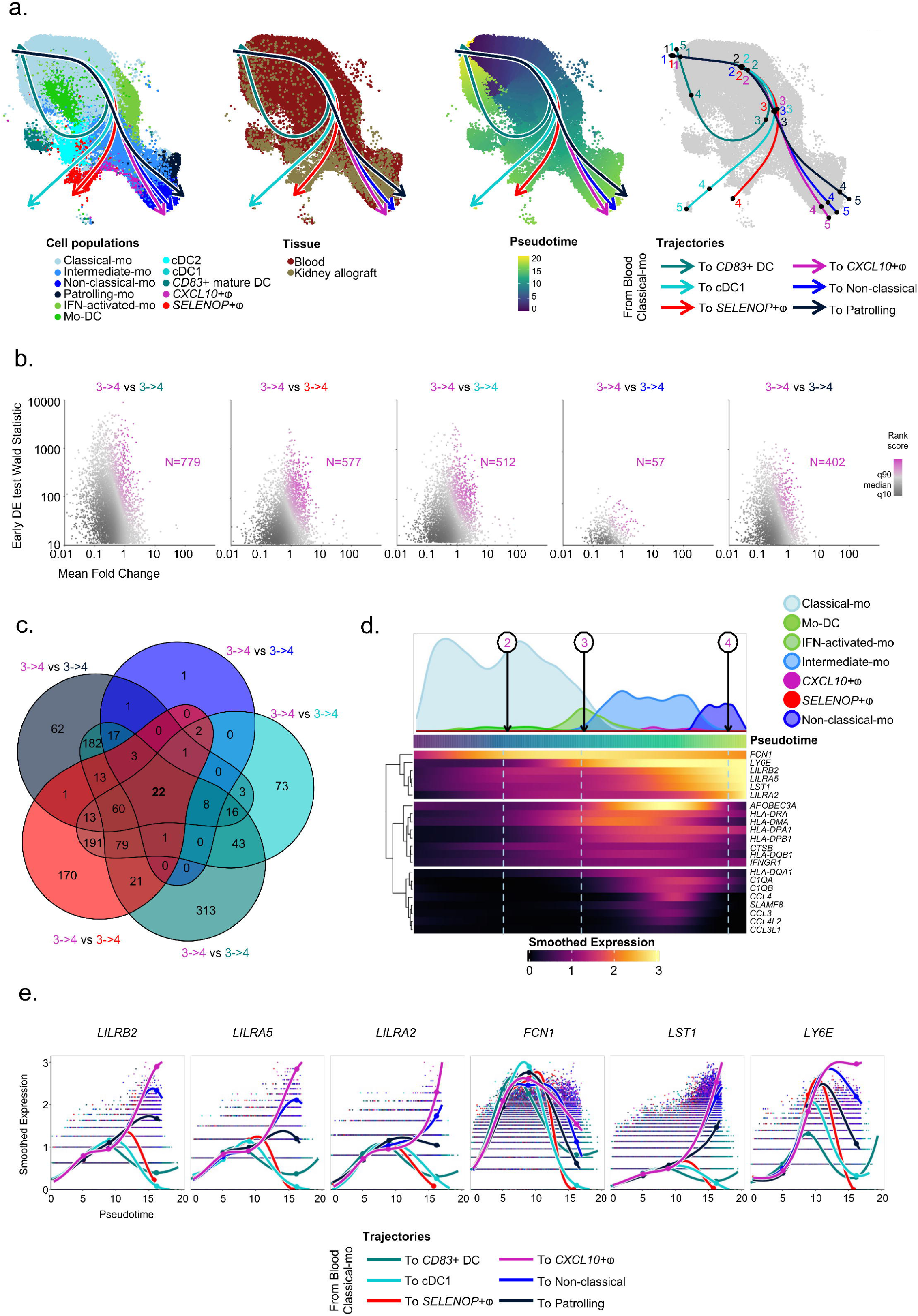
Myeloid cells differentiation is driven by six main trajectories in kidney transplant recipients. a. UMAP plots of reintegrated myeloid cells showing the 6 unsupervised trajectories obtained from slingshot and represented on cell type annotations and tissue origin, as well as pseudotime values corresponding to these trajectories and knots determined by tradeSeq and segmenting each lineage. b. Scatterplots representing early DE test between knot 3 and 4 computed statistics (Wald statistic, mean log fold change and rank score for each gene) of fitted GAM expression smoothers in 5 within-lineage comparisons (CD83+ DC trajectory versus *CXCL10*+L trajectory, cDC1 trajectory versus *CXCL10*+L trajectory, *SELENOP*+L trajectory versus CXCL10+L trajectory, Non-classical trajectory versus *CXCL10*+L trajectory and Patrolling trajectory versus CXCL10+L trajectory). c. Venn diagram showing overlapping genes among the top 90th percentile rank score in each of the 5 within-lineage comparisons of the *CXCL10*+ macrophages trajectory versus each other trajectory. d. Heatmap of the fitted GAM expression smoothers of 22 genes found to be significantly enriched in all 5 within-lineage comparisons of the *CXCL10*+ macrophages trajectory versus each other trajectory, shown alongside the pseudotime of the *CXCL10*+ macrophages lineage. Density of each cell type contributing to the trajectory is shown. e. Scatterplots of the log normalized expression and the corresponding fitted GAM expression smoothers curves of 6 significantly enriched genes in all 5 within-lineage comparisons of the *CXCL10*+ macrophages trajectory versus each other trajectory, represented alongside the pseudotime of all 6 identified lineages.

We then investigated whether this activated state due to the allogeneic responses could be captured in the blood of kidney transplant recipients. For this purpose, we built an ancillary cohort to the ORLY-EST cohort of kidney transplant recipients*^74^*. For each patient, a blood sample was collected on the day of transplantation (d0) and one year later (d365) (Fig. 6a). We therefore gathered N=64 patients with a pair of available PBMC samples (Fig. S6a). Of these patients, 8 showed mild to very severe rejection during the first year of transplantation (Fig. S6b). In order to perform a cytometric analysis limiting the potential batch effect, we followed a robust pipeline including batch correction and normalization (Fig. S6c). We then applied a gating strategy using CD14 and CD16 markers to select CD14+CD16-classical monocytes, CD14+CD16+ intermediate monocytes and CD14-CD16+ non-classical monocytes (Fig. S6d). By comparing the kinetics of LILRA1, LILRA2, LILRA5, LILRB1 LILRB2 and LILRB3 expression in these 3 populations between before transplantation and one year after, we observed that LILRA2 (GeoMFI_d0_ = 509 vs GeoMFI_d365_ = 626, P=0.046), LILRB1 (GeoMFI_d0_ = 565 vs GeoMFI_d365_ = 699, P=0.035) and LILRB2 (GeoMFI_d0_ = 1369 vs GeoMFI_d365_ = 1737, P=0.032) expression increases significantly in classical monocytes from patients with rejection compared with those without rejection. Moreover, this increase was not observed in intermediate or non-classical monocytes (Fig. 6b). Altogether, these results suggest that upon rejection, classical monocytes could differentiate into *CXCL10*+ macrophages and infiltrate the allograft or recirculate with the imprint of allogeneic activation.

**Fig. 6.**
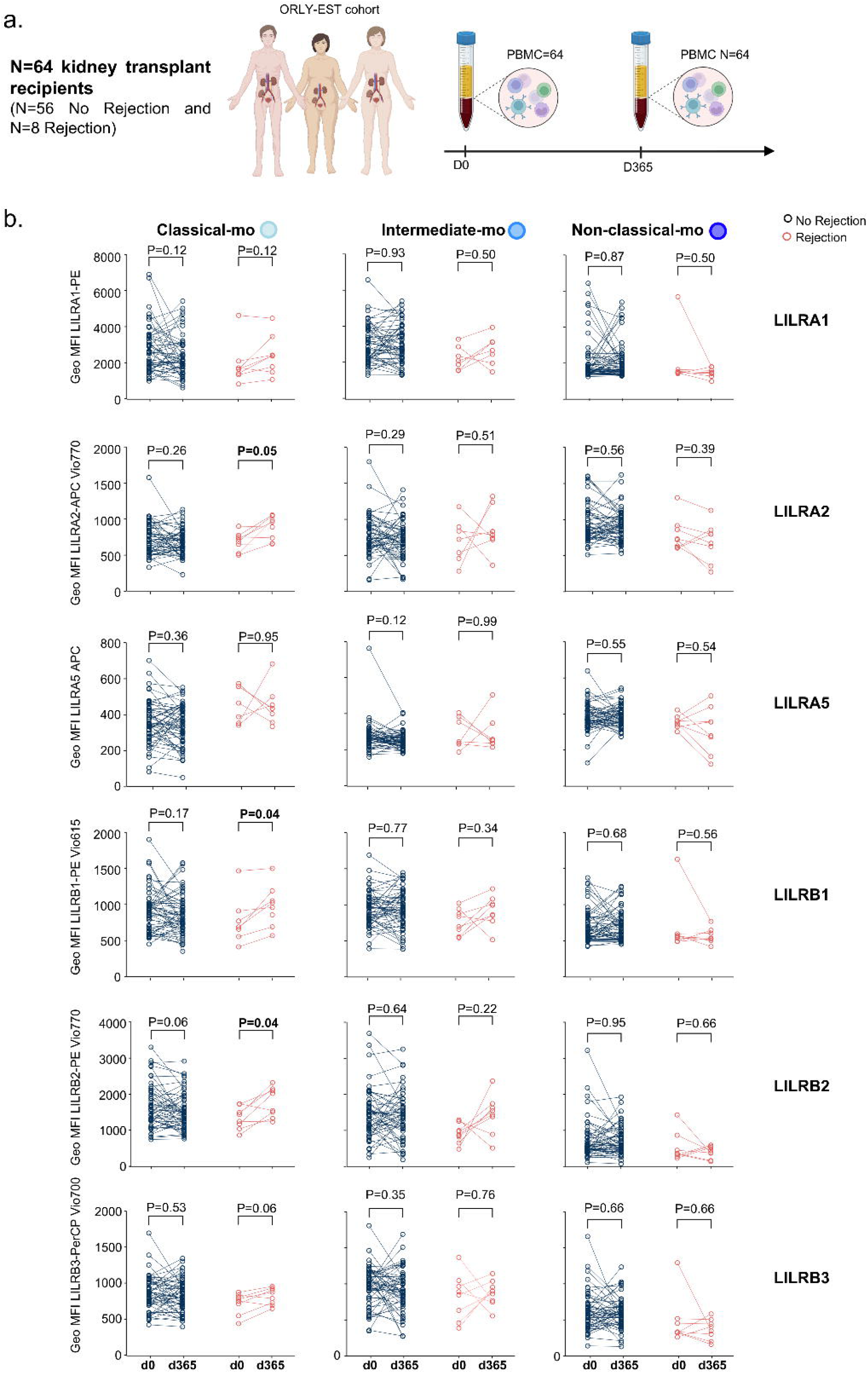
LILRA2, LILRB1 and LILRB2 expression increases significantly in classical monocytes from patients with rejection. a. Eighty-four patients were included in the ORLY EST multicenter cohort. PBMC and serum were collected at D0 and D365 post-transplant and stored at the CRB and the histocompatibility laboratory of Dijon University Hospital respectively. The proposed study is an ancillary study based on flow cytometric analysis of LILR expression in leukocytes. b. Normalized Expression of indicated markers at indicated times in indicated monocyte population. Multiple paired t tests were performed using Holm-Šídák method to compare the groups. P-values are shown.

### Allogeneic activation of classical monocytes leads to increased expression of LILRB2

Recent data from allogeneic recognition models in mice suggest that allogeneic monocytes are activated via the CD47-SIRP-⍰ axis*^71^*. We hypothesized that this activation could drive the differentiation of recipient-derived monocytes into *CXCL10*+ macrophages. To recapitulate the impact of the allogeneic activation in circulating classical monocytes within the allograft, we thus activated them *in vitro* by the CD47-SIRP-⍰ axis. For this purpose, classical monocytes were isolated from the blood of healthy volunteers and an immunological synapse was artificially mimicked as is widely and historically done for T cells by activating their CD3 and CD28 receptors with antibodies coated onto the culture plastic*^75^*. Here, we used polyclonal antibodies targeting CD47, essential for myeloid allo-activation*^10^* and measured the impact of these activations at protein levels after 48h of culture (Fig. 7a).

**Fig. 7.**
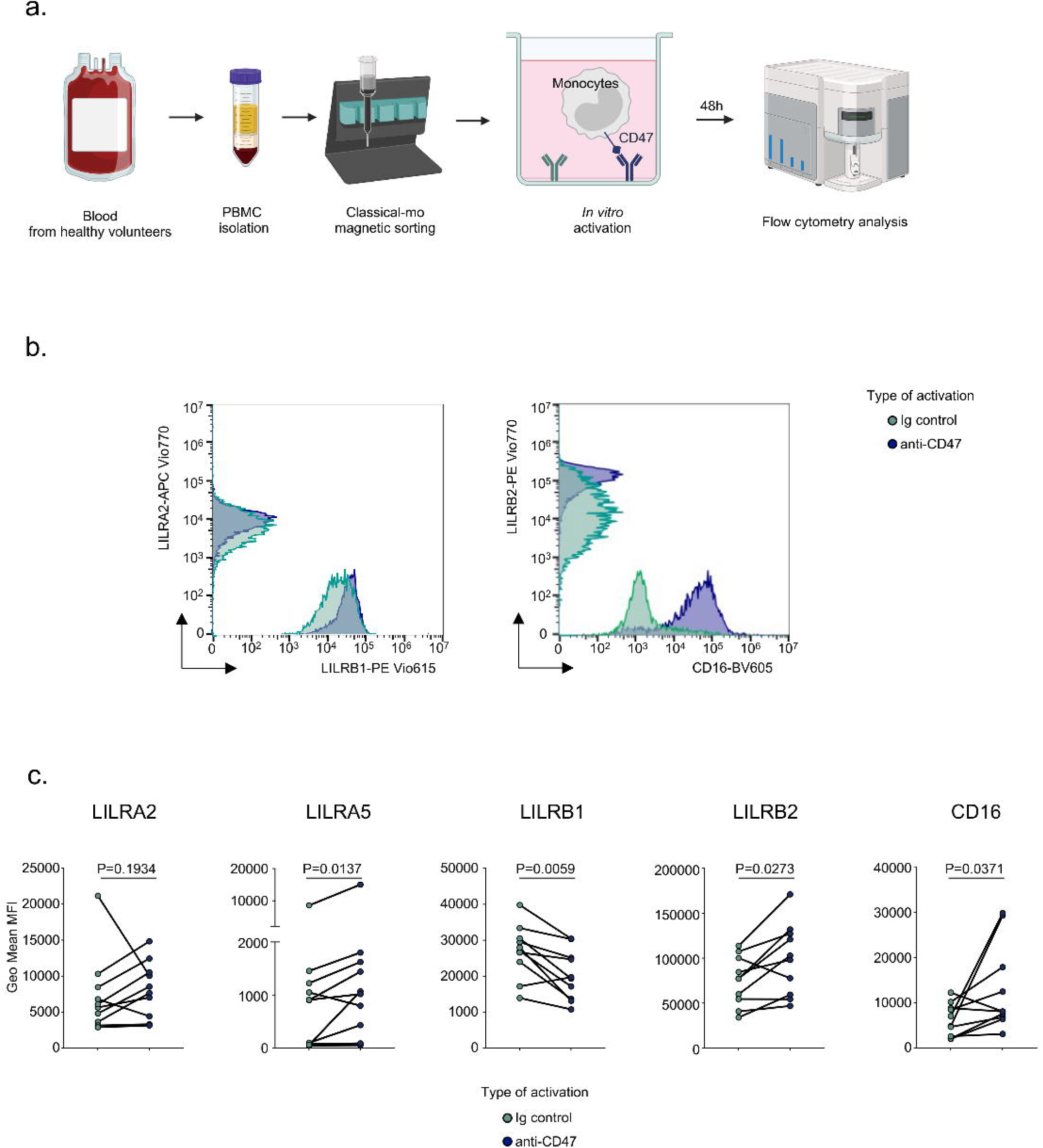
Allogeneic activation of classical monocytes leads to increased expression of LILRB2. a. Schematic diagram of the experiments. b. Representative overlays histograms representing the expression of annotated markers in different stimulatory conditions. c. Geometric Mean MFI of each biological replicate (N=10). Wilcoxon matched-pairs signed rank tests were used and P-values are shown.

Forty-eight hours after activation, LILRA2 was not differentially expressed at cell surface. In contrast, LILRB2 and LILRA5 were significantly increased after stimulation of CD47 (GeoMFI_Ig_ = 74975 vs GeoMFI_CD47_ = 99030, P=0.0273 for LILRB2 and GeoMFI_Ig_ = 1439 vs GeoMFI_CD47_ = 2338, P=0.0137 for LILRA5) compared with control condition. The expression of CD16 (GeoMFI_Ig_ = 6314 vs GeoMFI_CD47_ = 12952, P=0. 0371) was also doubled after CD47 stimulation. Surprisingly, LILRB1 expression was significantly decreased (GeoMFI_Ig_ = 26987 vs GeoMFI_CD47_ = 20437, P=0.0059, Fig. 5B-C). Altogether these results suggest that the allogeneic recognition of SIRP-17 via CD47 induces specifically certain LILRs such as LILRB2 and the differentiation from classical CD14+CD16-to intermediate CD14+CD16+ phenotype before macrophage differentiation.

Of note, SIRPA expression was primarily reported in myeloid cells but could also be detected in renal cells such as podocytes*^76–78^*. In addition, our cell-cell contact analysis suggested that CD47-SIRP-17 signaling in *CXCL10*+ macrophages relies on *SIRPA* expression in both *SELENOP*+ macrophages and PT_injured cells (Fig. S2d). We thus assessed the expression of SIRPA across all renal cell types and we confirmed that *SIRPA* is expressed in both podocytes and injured PT cells notably upon rejection (Fig. S7). These results suggest that *CXCL10*+ macrophages could continue to be activated through CD47-SIRP-17 in the tubulointerstitial compartment or in the damaged glomeruli within the allograft in addition to LILRs ligation.

### Macrophages overexpressing LILRB2 bind HLA class I molecules, triggering their activation

We then wanted to explore the impact of LILRB2 overexpression in macrophages. To do this, we started with a human myeloid cell line, the THP-1 cell line*^79^*, which, according to a previous report, lost LILRB2 expression on its surface compared with primary human monocytes*^80^*. We set up a transduction strategy with lentiviral vector encoding *LILRB2* and a reported gene, *TagBFP2* (Fig. S8a). We then successfully isolated TagBFP2+ single cell clones by FACS and therefore generated 2 clones, D02 and G02, of THP-1 expressing high level of LILRB2 (Fig. S8b). As LILRB2 is known to bind to class HLA molecules present on the cell surface, unlike LILRA2 or LILRA5 *^81,82^*, we next assessed whether LILRB2 overexpression could lead to a modulation of binding with HLA. Historically, THP-1 were described as HLA-A2+ and HLA-B5+*^79^*. We therefore verified that THP-1 express HLA-A2 and not HLA-B27 (Fig. S8c) and we then tested the LILRB2-MHC interaction in THP-1 WT and clones D02 and G02. In THP-1 WT that do not express LILRB2, binding to self HLA (assessed by HLA-A2 pentamers) or non self HLA (HLA-B27 pentamers) is undetectable. In contrast, in LILRB2 overexpressing clones D02 and G02, we detected fixation of both self and non-self HLA (Fig. S8d).

One of the limitations of the THP-1 model is inherent in the malignant nature of this cell line. We thus assessed the capacity of LILRB2-HLA interactions in *bona fide* primary human macrophages. In this aim, we adapted a similar lentiviral strategy in freshly isolated monocytes (Fig. 8a). After 10 days of M0 differentiation, we measured the expression of both TagBFP2 and LILRB2 in the transduced macrophages and observed a significant increase of the percentage of LILRB2hi cells (Fig. 8b-c) compared with non-transduced control (Median percent_NT_ = 0.00 vs Median percent_LILRB2_ = 53.90; P=0.0022). We applied the same LILRB2-HLA interactions assay using HLA-A2 and HLA-B27 pentamers and were able to detect class I HLA binding on LILRB2hi macrophages (Fig. 8d). We confirmed that LILRB2 overexpression on the surface of primary macrophage binds both to HLA-A2 (Median percent_NT_ = 1.09 vs Median percent_LILRB2_ = 3.60; P=0.0078, Fig. 8e) and HLA-B27 (Median percent_NT_ = 16.61 vs Median percent_LILRB2_ = 31.32; P=0.0273, Fig. 8f) suggesting that LILRB2 expression on the macrophages surface can be part of the immunological synapse between recipient-derived pro-inflammatory *CXCL10*+ macrophages and donor cells.

**Fig. 8.**
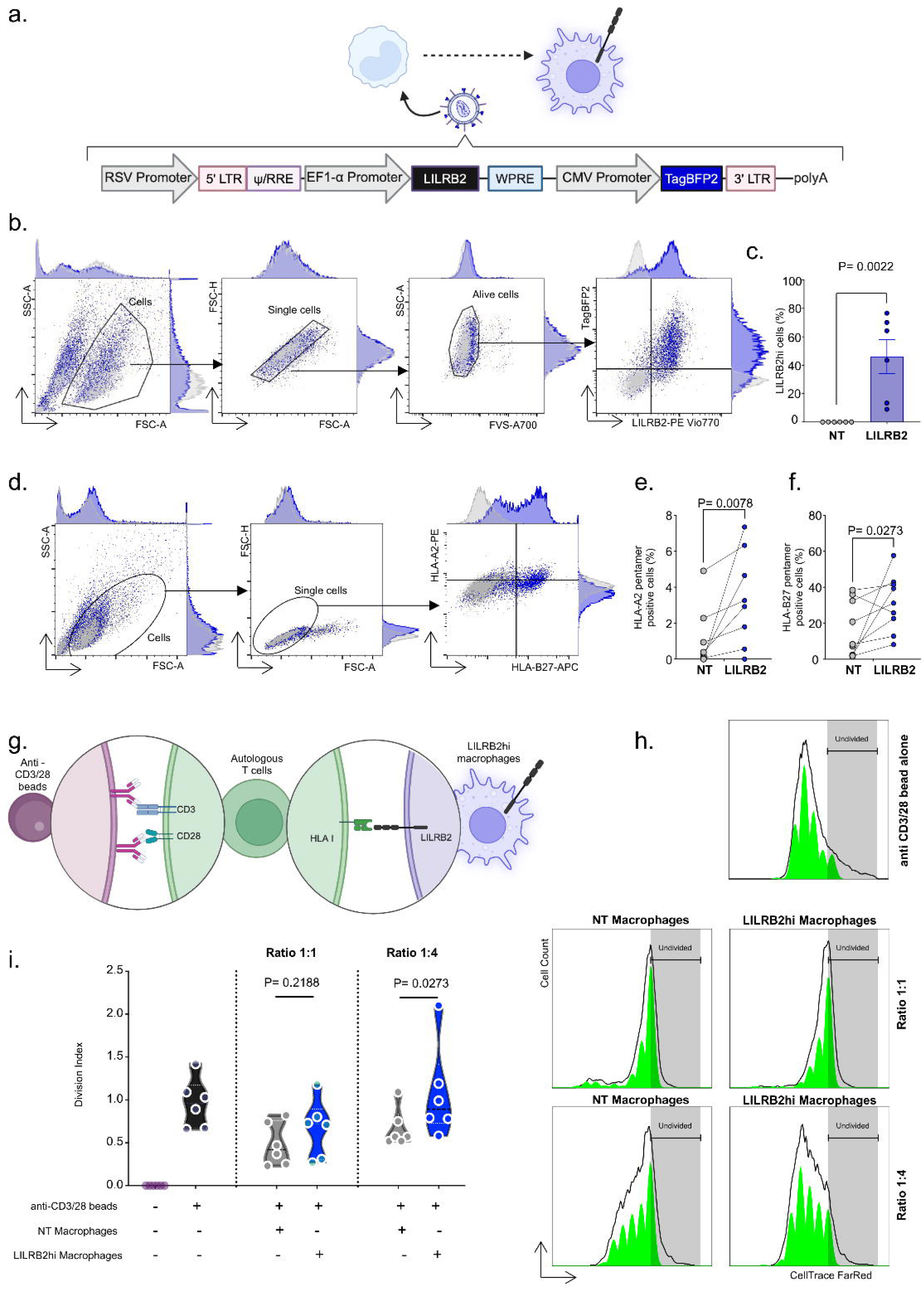
Macrophages overexpressing LILRB2 bind HLA class I molecules, triggering their activation. a. Schematic diagram of the lentiviral strategy used here. b. Gating strategy showing the efficacy of the approach. c. Percent of LILRB2 hi macrophages after transduction. Two-tailed Mann Whitney test was performed to compare the groups. P-values is shown. d.Gating strategy for HLA binding assays. e-f. Percentages of HLA-A2 or B-27 cells. Two-tailed Mann Whitney tests were performed to compare the groups. P-values are shown. g. Schematic diagram of CD3-CD28 activated autologous T cell proliferation assay upon co-culture with LILRB2 hi macrophages. h-i. Division index of the activated T cell post co culture with LILRB2 hi macrophages. Wilcoxon matched-pairs signed rank tests were performed to compare the NT and LILRB2hi conditions. P-values are shown.

A previous in vitro study has demonstrated that when LILRB2 is expressed on the surface of tolerogenic dendritic cells, it generates an antiproliferative signal to autologous T cells*^83^*. Here, we tested whether macrophages overexpressing LILRB2 would have an impact on the proliferation of autologous T cells (Fig. 8g). Compared with activated T cells alone, the addition of M0 macrophages to the culture slightly reduced cell proliferation, potentially through competition for nutrients (Fig. 8h). Interestingly, co-culture of LILRB2-overexpressing macrophages with T cells did not decrease T cell proliferation at a 1_macro_:1_T cell ratio_. Moreover, at a 1_macro_:4_T cell ratio_, T-cell proliferation increased significantly in the presence of macrophages overexpressing LILRB2 (P=0.0273, Fig. 6I) suggesting that LILRB2 overexpression at macrophages surface can generate a signal that positively impacts autologous T cell proliferation (Fig.9).

**Fig. 9.**
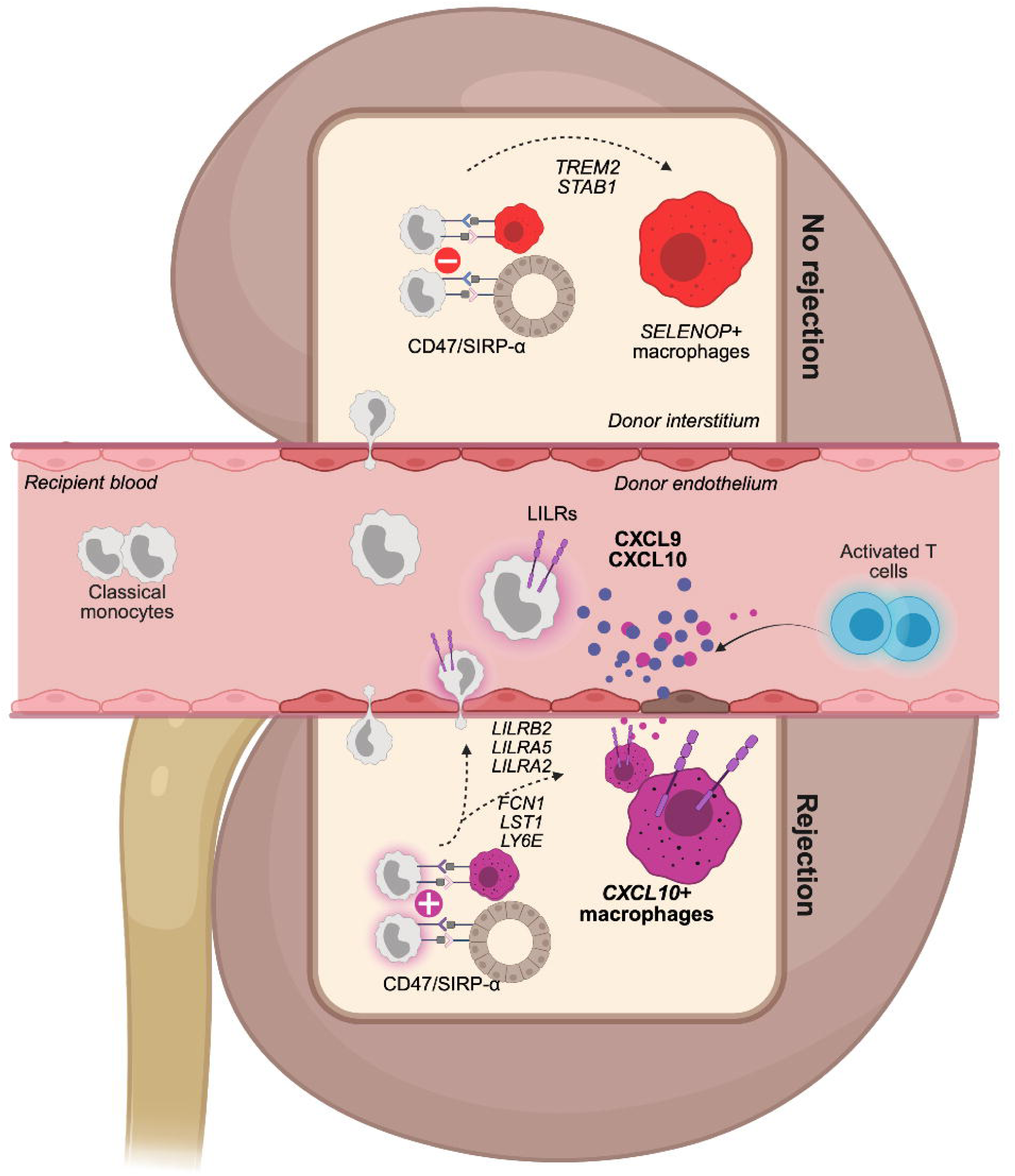
Mechanism of activation of kidney transplant recipient-derived monocytes in the context of kidney transplantation. The upper part of the figure suggests that in a context of balanced signals provided by CD47 and SIRPa, recipient-derived monocytes transform into *SELENOP*+ resident macrophages. In contrast, in the presence of an imbalanced CD47/SIRPA situation, monocytes transform into inflammatory macrophages expressing LILRB2 and CXCL10 or into memory monocytes overexpressing LILRB2.

## DISCUSSION

In the present study, we investigated circulating and allograft-infiltrating myeloid populations after kidney transplantation. To this end, we gathered a substantial amount of scRNAseq data from several study groups, several tissues (blood and biopsies) and several types of rejection in order to obtain a generalizable characterization of recipient monocyte/macrophage infiltration into the graft. Among the macrophage found within the allograft, we distinguished unsupervisedly two main populations, *CXCL10*+ macrophages and *SELENOP*+ macrophages. In addition to rejection-associated *CXCL10*+ macrophages, we identified *SELENOP*+ macrophages as the main macrophagic subset populating the kidney allograft. *SELENOP*+ macrophages show a very distinct transcriptomic profile as compare to *in vitro* differentiated M2 macrophages confirming that the M1-M2 classification is irrelevant in the context of allotransplantation and the description of macrophages *in vivo*. In line with our findings, Stewart and colleagues reported the existence of a subset of *bona fide* resident macrophages they annotated “MNP-d” and expressing high level of *SELENOP* (often also designated as *SEPP1*), *C1QC* and *RNASE1* in the native kidney*^84^*. In addition, these *SELENOP*+ macrophages specifically express *CD81*, a marker for resident macrophages that is conserved in mammalian species and more specifically in mouse, rat, pig and human kidney tissue*^85^*. Intriguingly, among *SELENOP*+ macrophages, only recipient-derived macrophages resulting from differentiation and infiltration of circulating monocytes expressed the trigger receptor expressed on myeloid cells 2 (TREM2). TREM2 was first described in macrophage infiltrating adipose tissue^86^, a population derived from monocytes and named “lipid-associated macrophages, LAMs” because they share a common gene expression program enriched in lipid metabolism-related genes, including *TREM2*, *APOE*, and *GPNMB^87^*. Here, we showed that donor-derived macrophages expressed high levels of *SELENOP* but not *TREM2* suggesting that TREM2 characterizes monocyte-derived macrophages that repopulate the kidney allograft after transplantation whereas *SELENOPhi* macrophages could derive from self-renewal of donor macrophages.

In contrast, *CXCL10*+ macrophages are uniquely derived from recipient monocytes and are major contributors of allograft inflammation during rejection independent of other immune cells. Inparticular, this population is absent in the allograft pre-transplantation and dramatically increases during rejection, expressing massive levels of the proinflammatory chemokines *CXCL10* and *CXCL9*. Importantly, these chemokines have been confirmed in a recent large-scale transcriptomic study as essential mediators of rejection in all types of solid organ transplantation, regardless of the transplanted organ or the subtypes of rejection*^3^*. Along with chemokine transcripts, *CXCL10*+ macrophages also strongly express *GBP1*, *GBP4*, *GBP5* and *WARS*, suggesting that these transcripts detected in bulk transcriptomic obtained from allograft biopsy samples and forming genes signatures of rejection*^62^* originate primarily from this macrophage population. Importantly, WARS and GBP1 were confirmed as enriched at protein level in the allograft upon rejection*^88,89^*. *CXCL10*+ macrophages also specifically express *ANKRD22* encoding Ankyrin repeat domain 22 (ANKRD22), a nuclear-encoded mitochondrial membrane protein that regulates mitochondrial Ca2+ and the Wnt pathway. *Ankrd22*^-/-^ mice showed reduced inflammation by inhibiting macrophages tissue infiltration and TNF secretion after gastric mucosal injury*^90^*. It is noteworthy that upregulation of ANKRD22 has previously been associated with macrophages activation in patients during kidney transplant rejection*^91^*.

At ontogenic level, *CXCL10*+ macrophages originate from recipient-derived circulating classical monocytes. These differentiate into intermediate monocytes, then progress either to a non-classical monocyte phenotype, as reported in the context of systemic infection*^73^*, or to macrophage differentiation. Importantly, we identified 6 core genes specifically associated with the *CXCL10*+ macrophages differentiation: *FCN1*, *LY6E*, *LST1*, *LILRA2*, *LILRB2* and *LILRA5*. Among these 6 genes, the expression of four of them, *FCN1*, *LST1*, *LILRA2* and *LILRB2*, is systematically increased in biopsies showing rejection, as shown by our external validations using the PROMAD atlas.

*FCN1* encodes Ficolin-1, a protein localized in the secretory granules of monocytes cytoplasm. Mechanistically, Ficolin-1 forms active proteolytic complexes with MBL-associated serine proteases, activating complement via the lectin pathway*^92^*. Importantly, C4b deposition increases with the amount of Ficolin-1*^93^*. Whether *CXCL10*+ macrophages can induce complement activation via Ficolin-1 sensing during kidney allograft rejection, independently of the humoral response remains to be investigated.

*LY6E* encodes a GPI-anchored cell surface protein called Lymphocyte antigen 6 complex locus E, which is involved in cell-cell adhesion and immunological synapse*^94^*. Further investigations must be pursued to determine the role of this protein in inflammatory macrophages infiltrating the graft.

*LST1* is encoded within the Major Histocompatibility Complex class III region on the short arm of chromosome 6, specifically including genes encoding components of the complement system such as C2, C4 and cytokines such as TNF*^95^*. *LST1* is notably included in the B-HOT panel as one of the transcripts most implicated in solid organ rejection*^63^*. It is highly polymorphic and its rs2256965 single nucleotide polymorphism is associated with a susceptibility for nephritis in systemic lupus erythematous patients*^96^*.*LILRB2*, *LILRA2* and *LILRA5* encode LILRs which have been shown to be essential in the allorecognition process of myeloid cells*^64,72^*. Like *LST1*, *LILRB2* is also included in the B-HOT panel *^63^*. Interestingly, cross-linking LILRA5 on the surface of monocyte induces the mobilization of calcium, a known signaling mediator that is released during cell activation. In addition, triggering of LILRA5 on monocytes induces the secretion of interleukin-1β (IL-1β), tumor necrosis factor-α (TNF-α), and IL-6. IL-1β, TNF-α, and IL-6 are normally released in the early stages of inflammatory responses, suggesting that LILRA5 might play a role in modulating monocyte function in inflammatory settings*^97^*.

While the ligand of LILRA5 and LILRA2 is still unknown*^82^*, LILRB2 has been reported as a putative binder for HLA class I*^98^*, which was previously confirmed *in vitro* in transfected 293T cells and ex vivo in CD14-myelomonocytic cells*^99^*. Here we showed that after CD47/SIRP-17 activation, classical monocytes overexpress LILRB2 on their surface together with CD16, suggesting that they initiate their differentiation into intermediate-monocytes. Moreover, this increase of LILRB2 in classical monocytes was also detected in the blood of patient with rejection one year after transplantation compared to patients with no rejection. Whether this increase in LILRB2 in circulating monocyte might reflect some myeloid immune memory towards nonself HLA in patients, as suggested in mice*^72^*, remains to be validated in larger cohorts in which SIRP-17 mismatches can be monitored. Interestingly, a population of proinflammatory macrophages strongly expressing LILRB2 has been reported to infiltrate the synovium of patients with rheumatoid arthritis*^100^*. LILRB2 was historically described as inhibitory because it includes ITIMs motifs at its intracellular part but it was recently reported that PirB/LILRB2 were expressed in hepatic macrophages and bound with their NASH-associated ligand ANGPTL8 to trigger the recruitment of macrophages to the liver, providing evidence that the LILRB2/PirB-ANGPTL8 axis could be a pathogenic driver of NASH pathogenesis and fibrogenesis. Notably, an PirB/LILRB2-ANGPTL8 interaction facilitates the differentiation of hepatic macrophages to a proinflammatory phenotype by enhancing the phosphorylation of P38, AKT, and P65 signals, which in turn causes an aggravation in hepatocyte lipid accumulation and an exacerbation from simple hepatic steatosis to steatohepatitis. This report also uncovers a function of PirB/LILRB2 receptors in monocytes migration which is in line with the present study*^101^* showing a specific increase in LILRB2 expression in recipient monocytes/macrophages infiltrating the allograft. Of note, ANGPTL8 is liver-specific and the LILRB2 ligand in kidney transplantation context remains to be characterized. Here, we showed that LILRB2 overexpression at macrophage’s surface can bind class I HLA and does not induce any antiproliferative signal to autologous T cells, suggesting that the increase of LILRB2 at *CXCL10*+ macrophages cell surface does not limit recipient T cells response during rejection. Instead, our results suggest that LILRB2, expressed by monocytes and then *CXCL10*+ macrophages, delivers an inflammatory signal to the myeloid cell in the allogeneic context. Our study does not allow us to conclude that it is the binding of LILRB2 to non-self HLA that induces this differentiation into *CXCL10*+ macrophages. For this, further investigations are required.

Our study has several limitations: we were unable to confirm the inflammatory monocyte/macrophage trajectories in another external dataset, but this work could be conducted in the coming years as the number of scRNASeq studies in human kidney transplantation increases every year. Spatial transcriptomics analyses would also allow us to better characterize the profile of *CXCL10*+ macrophages. It should be noted, that very recently, a spatial transcriptomics approach was carried out on FFPE-included renal graft biopsies, corroborating our findings. In this study, they confirm that monocytes and macrophages expressing *FCGR3A* are central to graft rejection, whatever the subtype of rejection. Using this new technique, they confirm that the *CXCL9*, *CXCL10*, *WARS*, *GBP1* and *GBP4* transcripts, which are *CXCL10*+ macrophages-specific transcripts as we described here, are universal transcripts associated with acute rejection. They also demonstrate upregulation of *CD47* and *SIRPA* in both TCMR and AMR suggesting that this CD47/SIRP-α axis is relevant in our study in order to investigate monocyte activation during allograft rejection*^102^*.

In addition, we were unable to integrate scRNASeq data from the blood of patients with non-AMR rejection and thus cannot conclude that blood monocytes behave in exactly the same way as in AMR or stable patients with this type of rejection. Nevertheless, given that intragraft chemokine secretion measurable in urine is increased in both AMR and TCMR, this may indicate that monocyte trajectories are similar in both types of rejection. Furthermore, our in vivo flow cytometry study of LILRs expression in patient’s blood is monocentric and limited by its small size. Moreover, it includes only a single post-transplant time point and does not allow us to speculate on the modulation of monocyte LILRB2 in the longer term. Larger-scale confirmations at several post-transplant timings are needed to demonstrate increased LILRB2 expression in classical monocytes, which may reflect allogeneic activation. Finally, our in vitro analyses on the impact of LILRB2 in inflammatory macrophages remains limited by the use of differentiation by LPS and IFN-γ which, as we have shown, does not 100% reflect the profile of *CXCL10*+ macrophages found within the graft. Of note, several teams have proposed and developed strategies to target monocytes and macrophages as a whole, with promising and encouraging results in the context of kidney transplantation *^103–105^*. The present study provides further insight into the pro-inflammatory axes of recipient-derived monocytes/macrophages, and suggests LILRB2 as a therapeutic target.

## Data Availability

All omic data are already publicly available.
All data produced in the present study are available upon reasonable request to the authors

## Acknowledgments

We thank the clinicians and surgeons, nursing staff and the patients of Dijon University Hospital. We thank Mickaël Ménager (Imagine Insitute, Paris) for providing the Vpx lentiviral vector. We thank Babacar Ndao (Inserm UMR Right, Besançon, France) for his help with fastq alignements. We thank Emilie Gaiffe, Caroline Laheurte and Eleonore Gravelin (Inserm UMR Right, Besançon, France) for their help with ORLY-EST samples. We thank Dr Christophe Masset (KU Leuven, Belgium) for providing the survival data associated with GSE147089. Figures 1a, 3a, 4a, 6a, 7a, 8a, 8g and 9 were created using Biorender.

## Data sharing statement

All omic data are already publicly available. All data produced in the present study are available upon reasonable request to the authors. The complete codebase developed for the unsupervised differentiation trajectory analysis of myeloid cells is publicly available as a Jupyter Notebook at https://github.com/InsermRightLab/LILRB2-Varin-et-al/. It provides a detailed, step-by-step walkthrough of the computational workflow used to generate each figure, encompassing pseudotime inference using slingshot and identification of differentially expressed genes via tradeSeq. The primary functions, especially those integral to the computational workflow, have been consolidated into RightOmicsTools, an R package available at https://alexis-varin.github.io/RightOmicsTools/. Additional supporting code, including utility functions, is provided within the notebook repository to ensure transparency and reproducibility. Code for Seurat is available at https://satijalab.org/seurat/. Code for CellChat is available at https://github.com/jinworks/CellChat/. Code for slingshot is available at https://github.com/kstreet13/slingshot/. Code for tradeSeq is available at https://github.com/statOmics/tradeSeq/.

