## Supplemental Material for "A subset of pro-inflammatory CXCL10+ LILRB2+ macrophages derives from recipient monocytes and drives renal allograft rejection"

Affiliations

### Table of Contents

### Supplementary Figures

**Supplementary Figure S1:** scRNAseq Quality control

**Supplementary Figure S2:** *In vitro* M1 vs M2 transcriptomic profiles, donor vs recipient macrophages proportion and CellChat analysis

**Supplementary Figure S3:** *CXCL10*+ macrophages are the myeloid cells most associated with allograft inflammation and rejection lesions

**Supplementary Figure S4:** PROMAD queries of the 6 core genes associated with *CXCL10*+ macrophages trajectory

**Supplementary Figure S5:** Expression of LILRs along pseudotime trajectories

**Supplementary Figure S6:** Quality control of flow cytometry analysis for the ORLY EST cohort

**Supplementary Figure S7** Expression of CD47 and SIRPA at single cell level

**Supplementary Figure S8:** LILRB2 induces HLA-A2 and HLA-B27 binding in THP1 cells

### Supplementary Tables

**Table S1** Specific markers used for cell annotations

**Table S2** Specific markers used for myeloid subset annotations

**SUPPLEMENTARY MATERIAL**

**Table S3** *CXCL10*+ and *SELENOP*+ macrophages gene signatures

### Supplementary References

### Supplementary Figures

### SUPPLEMENTARY MATERIAL

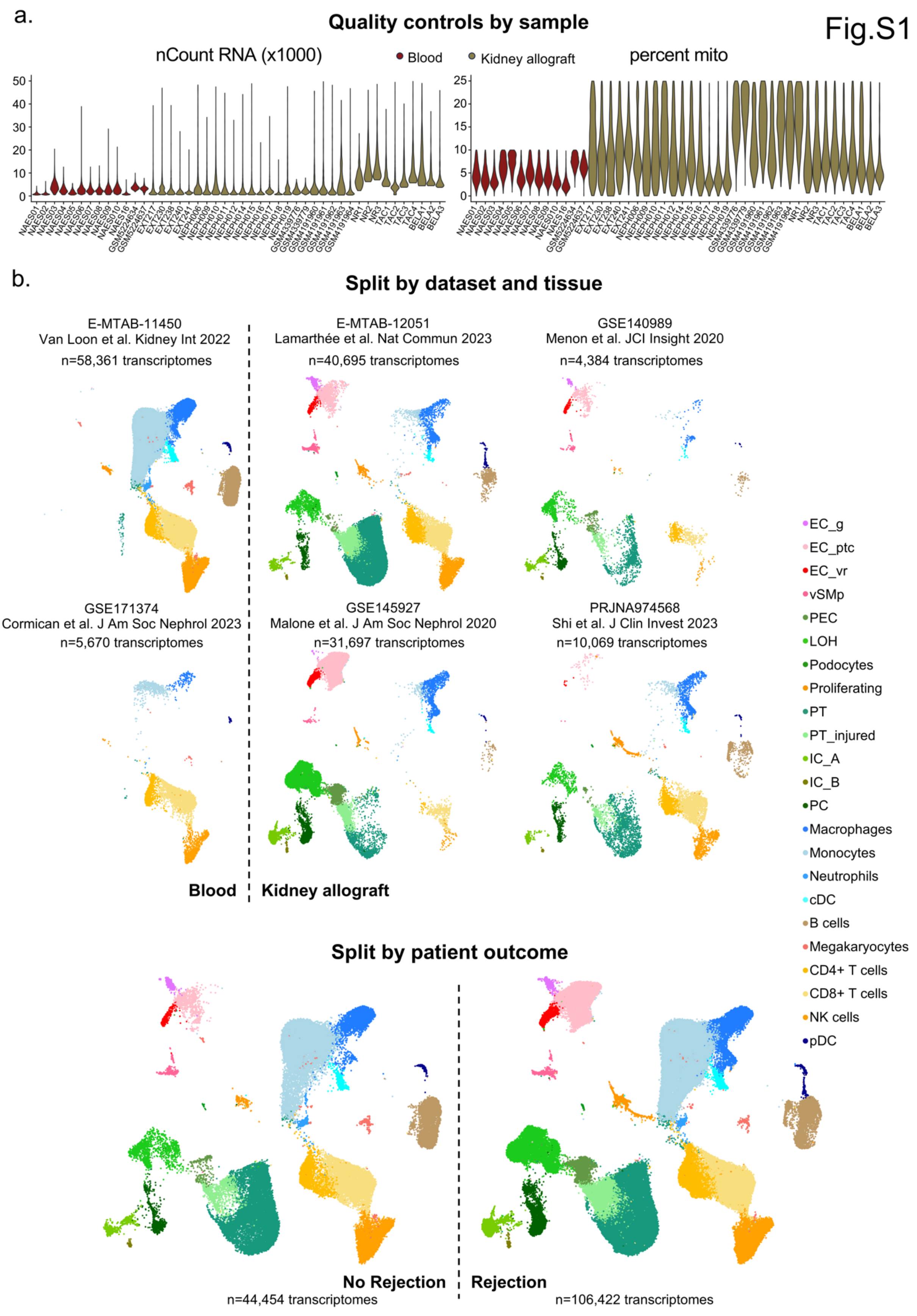

**SUPPLEMENTARY MATERIAL**

Figure S1. scRNAseq Quality control

a. Violin plots of the quality controls for each of the 46 samples, showing the sum of raw counts (nCount RNA) and the relative proportion of mitochondrial genes counts compared to total counts (percent mito).
b. Uniform Manifold Approximation and Projection (UMAP) plots of 150,876 cells passing QC filtering and doublets removal representing annotated cell types, split by dataset (2 from blood, E-MTAB-11450 and GSE171374, and 4 from kidney allograft biopsies, E-MTAB-12051, GSE140989, GSE145927 and PRJNA974568), or split by outcome (no rejection or rejection).

**SUPPLEMENTARY MATERIAL**

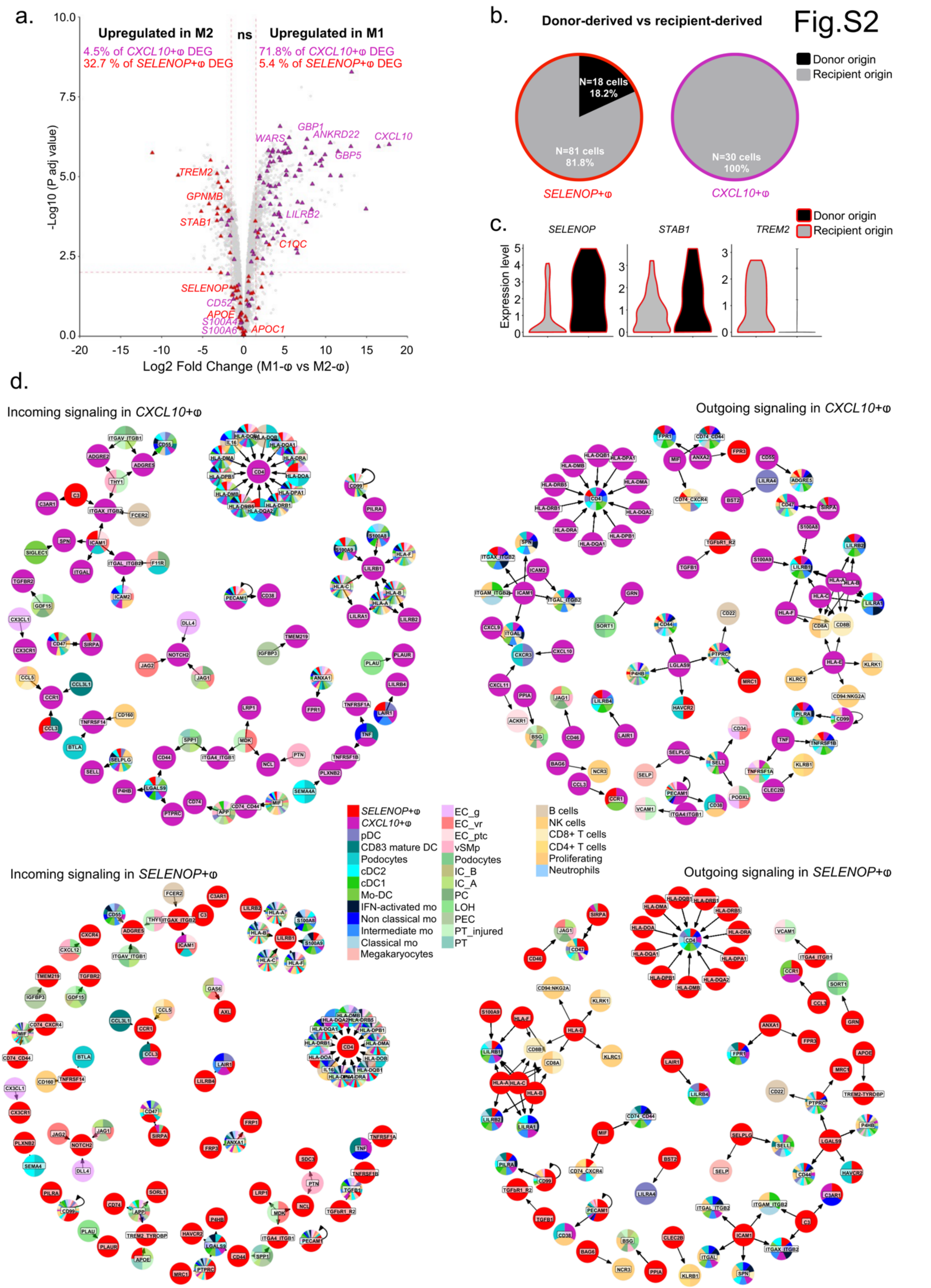


Figure S2. *In vitro* M1 vs M2 transcripomic profiles, donor vs recipient macrophages proportion and CellChat analysis

**SUPPLEMENTARY MATERIAL**

a. Volcano plot from an external validation bulk RNA-Seq dataset, GSE146028, representing DEG between M1 (N=3 donors) versus M2 macrophages (N=3 donors) differentiated *in vitro* from isolated monocytes, with specific markers found between *SELENOP*+ and *CXCL10*+ macrophages highlighted.
b. Pie charts showing the proportions of donor-derived and recipient-derived *SELENOP*+ and *CXCL10*+ macrophages identified in samples with a sex mismatch between the donor and the recipient.
c. Violin plots representing markers expression (log normalized) in donor-derived or recipient-derived *SELENOP*+ macrophages from samples with a sex mismatch between the donor and the recipient.
d. Networks showing incoming (signals received by the receptors) and outgoing (signals sent from the ligands) communications of respectively *SELENOP*+ and *CXCL10*+ macrophages with all other cell types, inferred by CellChat analysis.

SUPPLEMENTARY MATERIAL

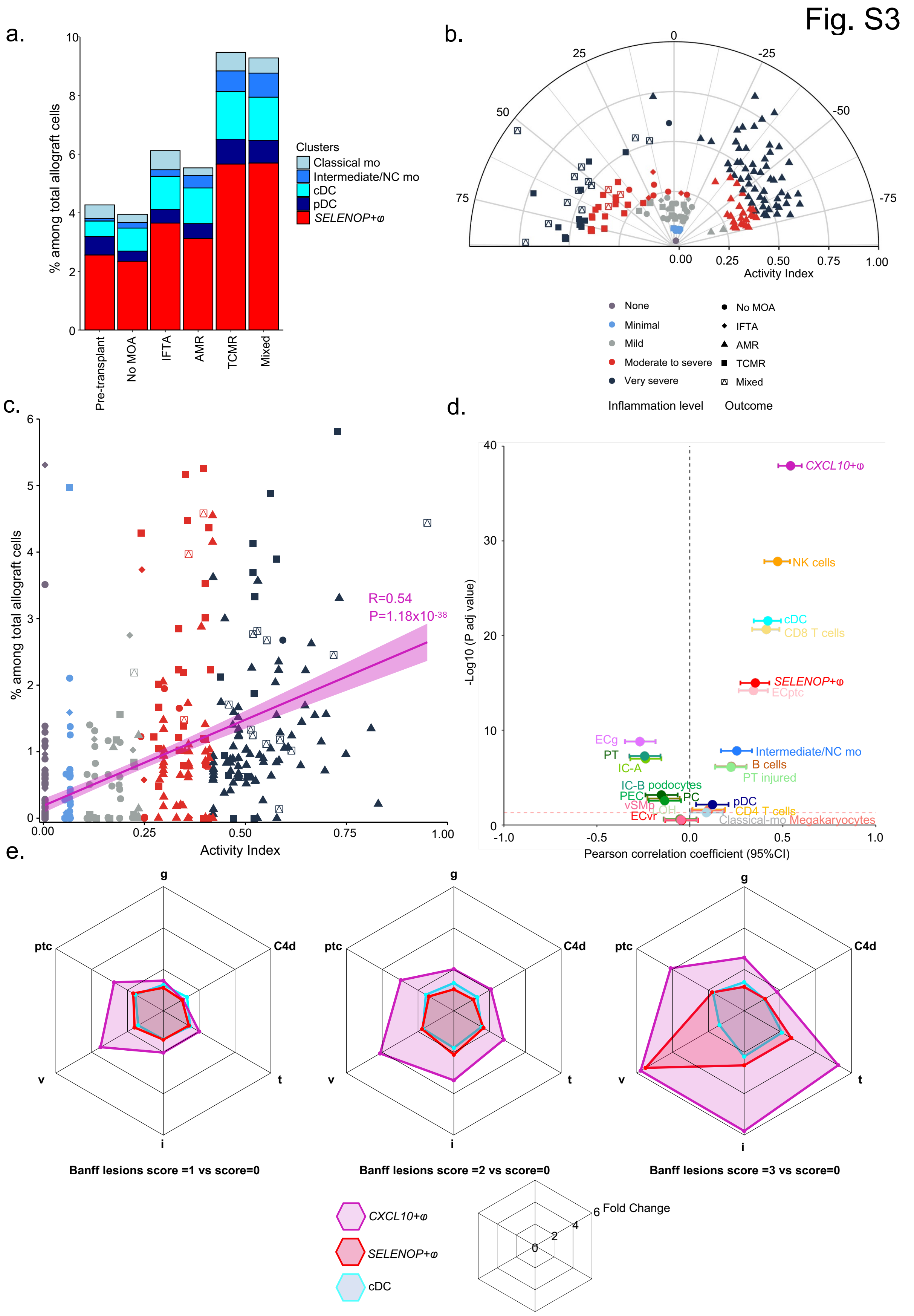


Figure S3: *CXCL10+* macrophages are the myeloid cells most associated with allograft inflammation and rejection lesions.

### SUPPLEMENTARY MATERIAL

a. Proportion of the other myeloid subsets (cDC, classical and non classical monocytes, pDC and *SELENOP*+ macrophages) in pre-transplant and post-transplant biopsies according to patient outcomes. b. Polar plot depicting the heterogeneity of histological profiles in GSE98320. c. Correlation between inflammation (Activity Index) and the proportion of *CXCL10*+ macrophages inferred by deconvolution in GSE98320. d. Pearson's correlation coefficient is represented by a dot, with the 95% confidence interval indicated by error bars, for the correlation between the Activity Index and the frequency of different immune cells within the allograft, inferred by deconvolution in GSE98320. e. Radar plots showing the fold change of the proportions of *CXCL10*+ macrophages, *SELENOP*+ macrophages and cDC according to the Banff scores of acute rejection lesions and compared to the score =0 condition.

Fig.S4

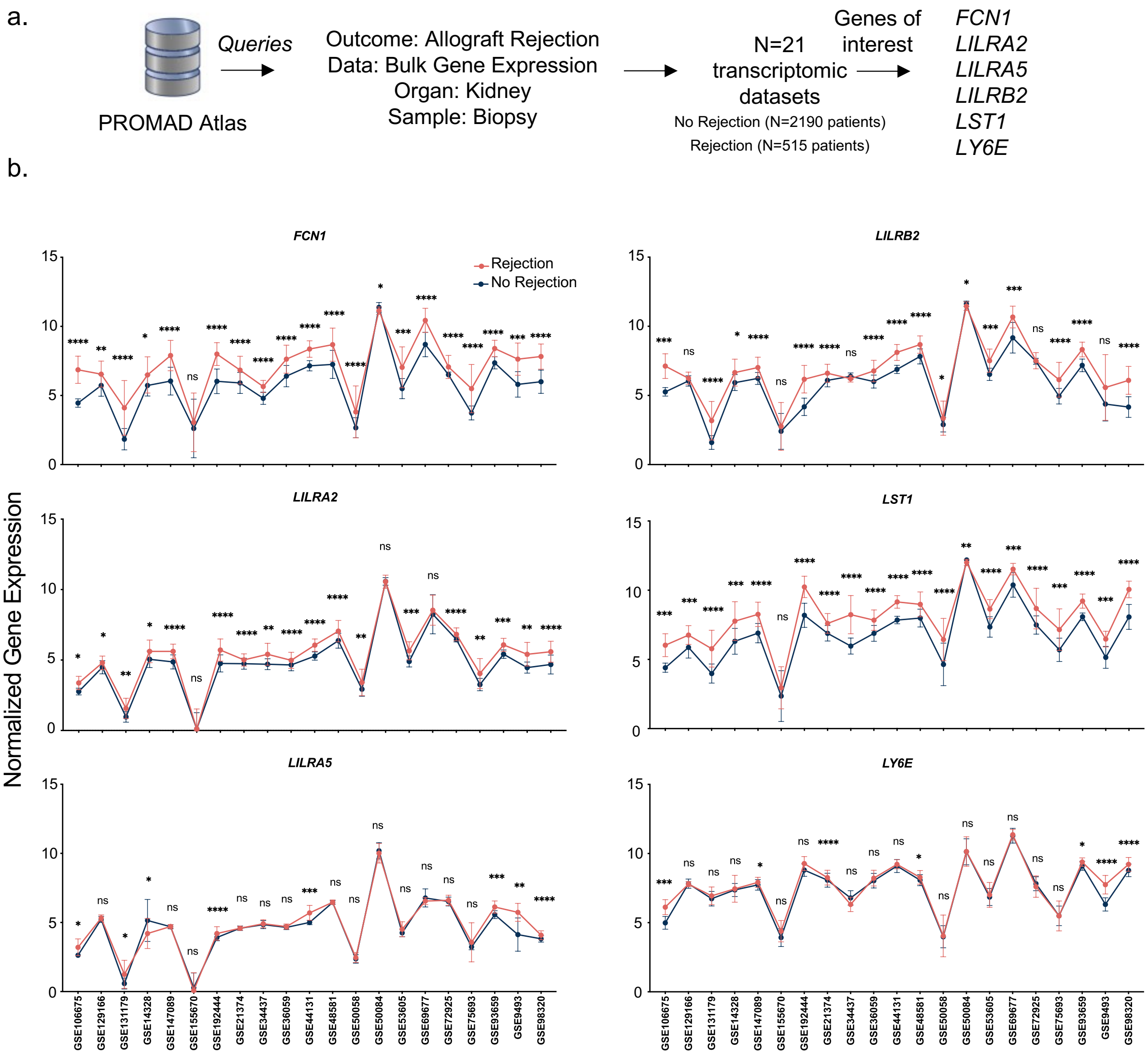


Figure S4. PROMAD queries of the 6 core genes associated with *CXCL10*+ macrophages trajectory

a. PROMAD querries. b. PROMAD results in external transcriptomic datasets including kidney allograft biopsies-derived relative expression of each annotated gene. Two-tailed unpaired t test was used to compare the expression between groups in each dataset. * p value>0.05, ** p value>0.01, *** p value>0.001, **** p value>0.0001,

Fig.S5

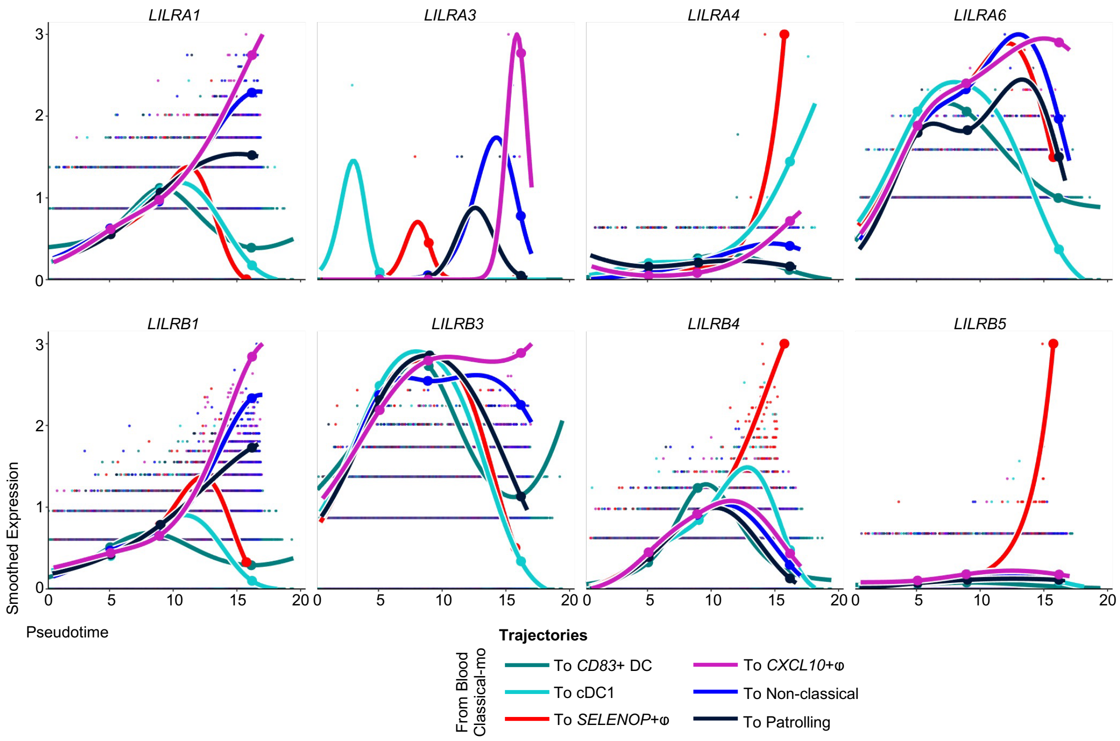


Figure S5. Expression of LILRs along pseudotime trajectories
Scatterplots of the log normalized expression and the corresponding fitted GAM expression smoothers curves of innate allorecognition markers, represented alongside the pseudotime of all 6 lineages identified by slingshot.

### SUPPLEMENTARY MATERIAL

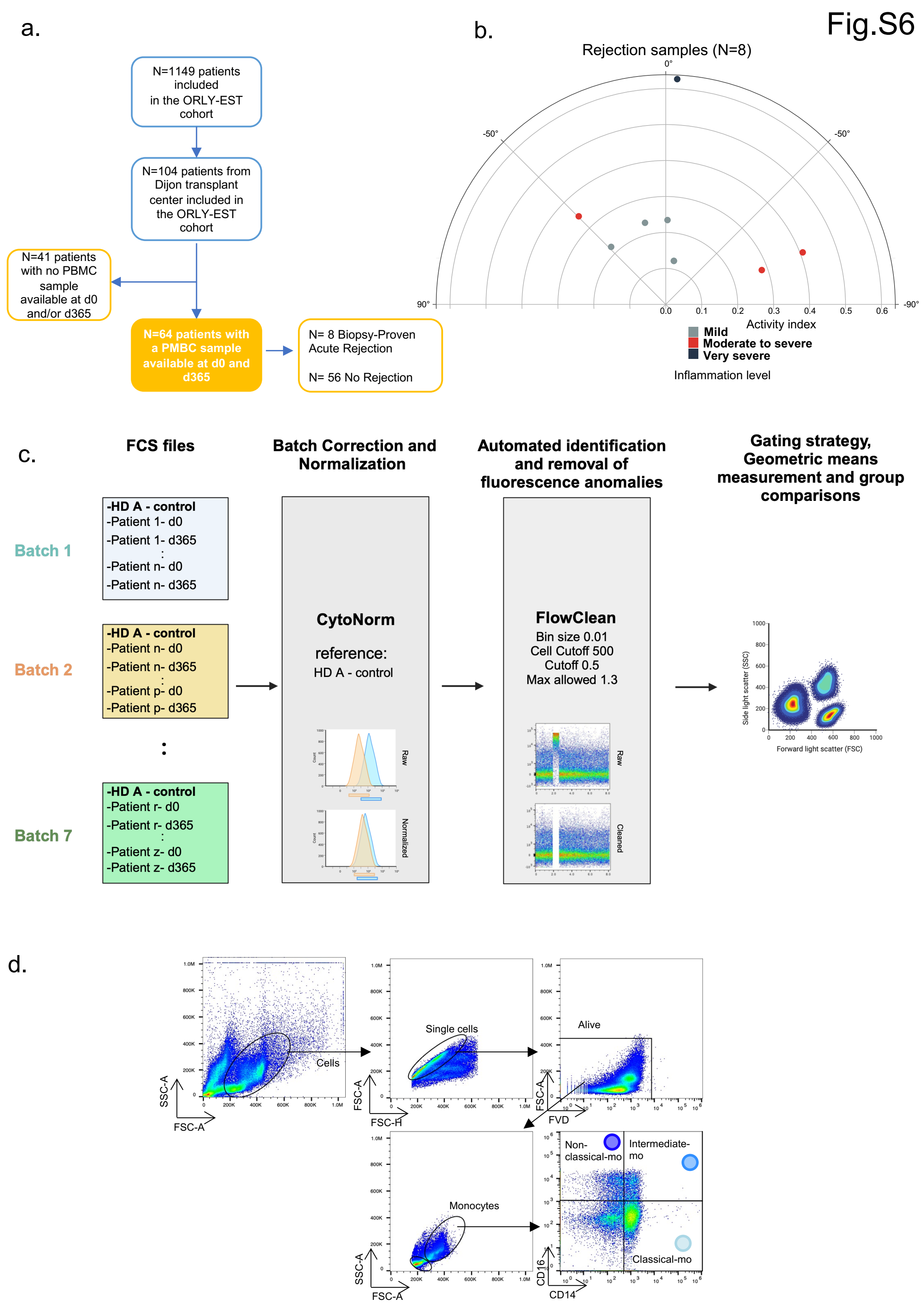


Figure S6. Quality control of flow cytometry analysis for the ORLY EST cohort

Of the 1150 patients included in the ORLY-Est cohort between 2008 and 2020, 107 were transplanted at Dijon University Hospital. To ensure sufficient cellular material for all the planned experiments, only patients with two PBMC samples available at d0 and d365 were included. Thus, 81 had samples available at both times, two did not at d0 and 24 did not at d365. Fourteen patients were excluded for not having at least two samples available at both times, and for 3 others, samples could not be found at the Biological Resource Center. A total of 64 patients were included in this ancillary study. To ensure analysis quality, cytometry files were normalized between runs using the CytoNorm[1] plugin, and abnormal events intrinsic to cytometry were removed using the FlowClean[2] plugin. Gating strategies excluded doublets and dead cells before selecting monocytes according to size (FSC-A) and granularity (SSC-A). Monocyte subpopulations were then identified using CD16 and CD14 markers in live total monocytes. Classical monocytes were defined as CD16-CD14+, intermediate monocytes as CD16+ CD14+ and non-classical monocytes as CD16+ CD14-. The normalized geometric mean fluorescence of LILRB2 was then measured in these three subpopulations.

Fig.S7

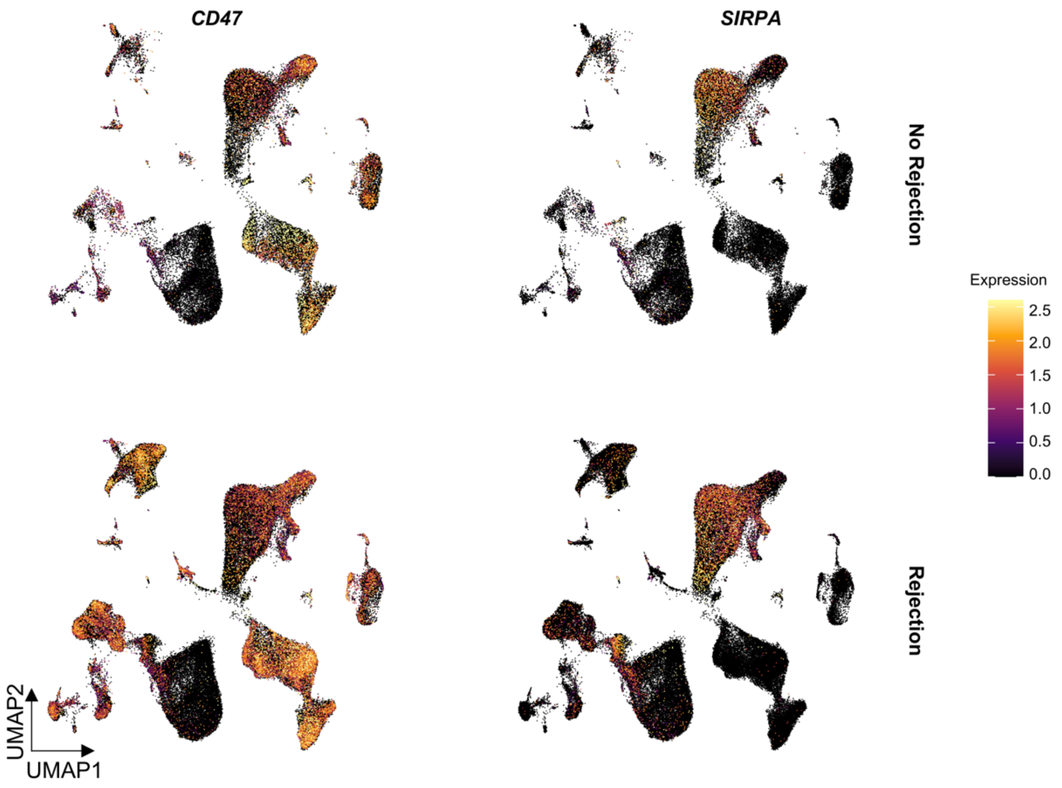


Figure S7. Expression of CD47 and SIRPA at single cell level
UMAP plots, split by outcome (no rejection or rejection), showing innate allorecognition markers expression (log normalized).

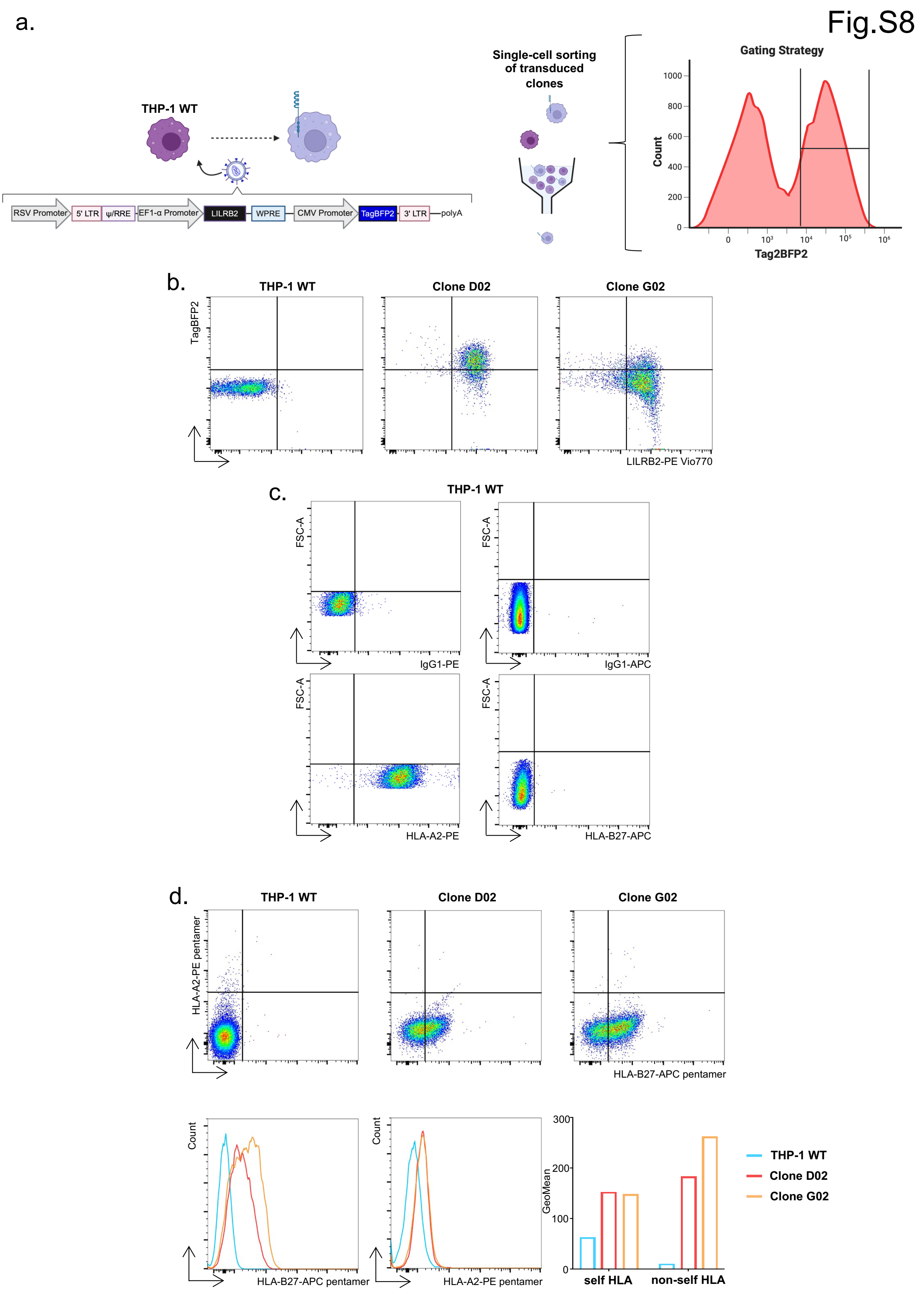


Figure S8. LILRB2 induces HLA-A2 and HLA-B27 binding in THP1 cells
a. Schematic strategy for THP1 transduction and clonage b. Dot plots representing the expression of LILRB2 and BFP2 in WT THP1 and in the two clones obtained after transduction. c. Dot plots

representing the HLA-A2 and B27 expression in THP1 cell line. d. Analysis of HLA pentamer binding on WT THP1 and the two transduced clones.

### Supplementary Tables

| Clusters | Annotation Markers | | |
|---|---|---|---|
| PT | ***SLC5A12***[3,4] | ***ALDOB***[5,6] | *LRP2*[3,7] |
| PT_injured | *VCAM1*[3,4,7] | *HAVCR1*[3,4] | ***CDH6***[3] |
| PEC | *CLDN1*[3,8–10] | *LIX1*[5,10] | ***CTGF***[8] |
| LOH | ***UMOD***[3–8] | ***SLC12A1***[3–5,8] | ***IRX2*** |
| PC | ***SCNN1G***[3,5,7,8] | ***AQP2***[3,5,8] | ***FXYD4***[3,5,8] |
| IC_A | ***DMRT2***[8] | ***SLC4A1***[3–5,7] | *SLC26A7*[3–5,8] |
| IC_B | ***SLC26A4***[5] | ***INSRR***[3,5] | ***HMX2***[5] |
| Podocytes | ***NPHS1*** | ***NPHS2***[3,4,8] | ***KLK7*** |
| vSMp | *ACTA2*[3–5,7,8] | *TAGLN*[3–5,8] | ***FRZB***[5] |
| EC_ptc | ***PLVAP***[3–5,8] | ***ACKR1***[5] | ***DNASE1L3***[3,5,8] |
| EC_vr | ***CLDN5***[4,5,8] | ***SOX17***[5] | ***FBLN2*** |
| EC_g | *EMCN*[3,4,8] | ***CRHBP***[5,8] | ***SOST***[5,8] |
| Megakaryocytes | *PPBP*[11,12] | *PF4*[13] | ***GP9***[5] |
| Proliferating | *MKI67*[3,6] | ***UBE2C*** | ***HJURP*** |
| CD4+ T cells | ***LEF1*** | ***IL7R***[3–5,7,8,13] | ***CD3G***[5,8,12] |
| CD8+ T cells | ***CD8A***[4,5,7,11–13] | ***GZMK***[8] | ***THEMIS***[3] |
| NK cells | ***GNLY***[3–5,7,8,11] | ***KLRF1*** | ***SH2D1B*** |
| B cells | *MS4A1*[3–5,7,8,11,12] | ***CD79A***[8,13] | ***CD19***[4–6,12] |
| Monocytes | ***S100A12***[5,14] | *CD14*[5,11–13,15] | ***LYZ***[5,11] |
| Macrophages | ***C1QA***[5] | ***C1QB***[5,7] | ***MS4A4A*** |
| cDC | ***CLEC10A***[4] | ***CD1C***[4,5,11,12] | ***FCER1A***[5,13] |
| pDC | *IL3RA*[3,5,11] | ***LILRA4***[12,13] | ***LRRC26*** |
| Neutrophils | ***CSF3R***[16] | *NAMPT*[16] | ***DMXL2*** |

Table S1: Specific markers used for cell annotations. Genes written in bold were among the top 5 most differentially expressed markers (as defined by highest log2 fold change among statistically significant FDR adjusted p-value genes) unsupervisedly associated with the indicated clusters, found by Seurat's FindMarkers function. The other genes are canonical markers from the indicated literature.

**SUPPLEMENTARY MATERIAL**

| Clusters | Myeloid Markers | | |
|---|---|---|---|
| Classical-mo | ***S100A12***[5,14] | *CD14*[5,11–13,15] | *LYZ*[5,11] |
| Intermediate-mo | *MARCO*[14] | ***APOBEC3A***[14] | ***HLA-DPB1***[14] |
| Non-classical-mo | ***FCGR3A***[4,5,11] | ***CDKN1C***[5,14] | ***NEURL1*** |
| Patrolling-mo | ***CX3CR1***[17] | ***PECAM1***[17] | ***TCF7L2*** |
| IFN-activated-mo | ***IFIT1***[18] | ***HERC5***[18] | ***MX1***[18] |
| Mo-DC | ***LMNA***[19,20] | ***AHNAK*** | ***HIST1H1E*** |
| cDC2 | ***CLEC10A***[4] | ***FCER1A***[5,13] | ***CD1C***[4,5,11,12] |
| cDC1 | ***XCR1***[4] | ***GCSAM*** | ***CLEC9A***[4,5] |
| *CD83*+ mature DC | ***TNF*** | ***CCL3L1*** | *CD83*[21] |
| *CXCL10*+φ | *C1QA*[5] | ***CXCL10*** | ***ANKRD22*** |
| *SELENOP*+φ | *STAB1*[7] | ***TREM2*** | ***SELENOP*** |

Table S2 Specific markers used for myeloid subset annotations. Genes written in bold were among the top 5 most differentially expressed markers (as defined by highest log2 fold change among statistically significant FDR adjusted p-value genes) unsupervisedly associated with the indicated clusters, found by Seurat's FindMarkers function. The other genes are canonical markers from the indicated literature.

| Gene | avg_log2FC | pct.*CXCL10*+ macrophage | pct.*SELENOP*+ macrophage | -Log10 (p value adj) | Signature | Presence in B-HOT panel |
|---|---|---|---|---|---|---|
| A2M | -4.909164069 | 0.022 | 0.469 | 62.06769004 | SELENOP+ macrophage | No |
| ADAM28 | -5.695026686 | 0.016 | 0.288 | 31.17036484 | SELENOP+ macrophage | No |
| ADGRE1 | 2.808276153 | 0.249 | 0.053 | 18.15041403 | CXCL10+ macrophage | No |
| ADGRE5 | 1.828339231 | 0.748 | 0.335 | 46.55327033 | CXCL10+ macrophage | No |
| AKR1B1 | -3.144123957 | 0.141 | 0.462 | 37.66238134 | SELENOP+ macrophage | No |
| ALOX5AP | -5.148537739 | 0.038 | 0.432 | 51.51505012 | SELENOP+ macrophage | No |
| ANKRD22 | 1.898205285 | 0.537 | 0.198 | 29.89621753 | CXCL10+ macrophage | Yes |
| APOBEC3A | 4.496881225 | 0.545 | 0.045 | 85.26708935 | CXCL10+ macrophage | No |
| APOC1 | -3.797702833 | 0.068 | 0.597 | 72.6118689 | SELENOP+ macrophage | No |
| APOE | -5.45808007 | 0.06 | 0.617 | 87.23130399 | SELENOP+ macrophage | Yes |
| APOL3 | 2.473302627 | 0.615 | 0.221 | 53.72832929 | CXCL10+ macrophage | No |
| APOL6 | 1.546757649 | 0.618 | 0.281 | 29.47986277 | CXCL10+ macrophage | No |
| ARAP2 | 2.267829855 | 0.371 | 0.121 | 22.16817261 | CXCL10+ macrophage | No |
| ARHGAP18 | -2.445018854 | 0.125 | 0.389 | 19.41273147 | SELENOP+ macrophage | No |
| AXL | -4.179930814 | 0.038 | 0.306 | 26.76476667 | SELENOP+ macrophage | Yes |
| BAZ1A | 1.746755677 | 0.593 | 0.283 | 33.31536576 | CXCL10+ macrophage | No |
| BCL2A1 | 1.742316847 | 0.659 | 0.274 | 43.77243313 | CXCL10+ macrophage | Yes |
| BHLHE41 | -3.441852513 | 0.024 | 0.228 | 16.71119354 | SELENOP+ macrophage | No |
| BID | 2.020073765 | 0.648 | 0.297 | 49.83729501 | CXCL10+ macrophage | No |
| BIRC3 | 2.30803798 | 0.474 | 0.115 | 36.16697637 | CXCL10+ macrophage | Yes |
| C15orf39 | 1.91272848 | 0.407 | 0.103 | 26.89278619 | CXCL10+ macrophage | No |
| C19orf38 | 2.081382808 | 0.431 | 0.158 | 24.75565444 | CXCL10+ macrophage | No |
| C1QC | -1.929937336 | 0.398 | 0.875 | 73.96519135 | SELENOP+ macrophage | No |
| C3 | -4.343573765 | 0.06 | 0.478 | 52.40281814 | SELENOP+ macrophage | Yes |
| CAPG | -1.626569412 | 0.341 | 0.513 | 17.62936228 | SELENOP+ macrophage | No |

**SUPPLEMENTARY MATERIAL**

| | | | | | | |
|---|---|---|---|---|---|---|
| CARD16 | 1.582592232 | 0.756 | 0.414 | 47.5865346 | CXCL10+ macrophage | Yes |
| CASP5 | 2.911041308 | 0.206 | 0.026 | 19.99711133 | CXCL10+ macrophage | No |
| CD163 | -1.985981433 | 0.198 | 0.448 | 15.81896988 | SELENOP+ macrophage | Yes |
| CD300E | 2.535102905 | 0.341 | 0.077 | 23.61321628 | CXCL10+ macrophage | No |
| CD48 | 1.682580391 | 0.734 | 0.349 | 47.70765571 | CXCL10+ macrophage | Yes |
| CD52 | 2.802931776 | 0.813 | 0.223 | 98.5841564 | CXCL10+ macrophage | No |
| CD81 | -1.94216968 | 0.22 | 0.479 | 19.34570407 | SELENOP+ macrophage | Yes |
| CD9 | -4.086513555 | 0.041 | 0.353 | 32.3404215 | SELENOP+ macrophage | No |
| CFP | 2.51599325 | 0.696 | 0.171 | 69.09094645 | CXCL10+ macrophage | No |
| CPPED1 | 1.765814597 | 0.48 | 0.204 | 20.96105194 | CXCL10+ macrophage | No |
| CREG1 | -1.771462591 | 0.249 | 0.449 | 16.61735023 | SELENOP+ macrophage | No |
| CRIP1 | 2.622431003 | 0.572 | 0.214 | 38.93830974 | CXCL10+ macrophage | No |
| CSF2RB | 1.751548278 | 0.388 | 0.132 | 18.6062696 | CXCL10+ macrophage | Yes |
| CTSD | -1.993318845 | 0.485 | 0.6 | 29.22307925 | SELENOP+ macrophage | No |
| CXCL10 | 3.919718164 | 0.808 | 0.16 | 116.4357704 | CXCL10+ macrophage | Yes |
| CXCL11 | 3.709248324 | 0.301 | 0.055 | 27.38829691 | CXCL10+ macrophage | Yes |
| DAB2 | -4.117591642 | 0.084 | 0.462 | 44.33718896 | SELENOP+ macrophage | No |
| DRAM1 | 1.560723716 | 0.52 | 0.222 | 23.00522913 | CXCL10+ macrophage | No |
| EHD1 | 2.137143666 | 0.355 | 0.098 | 21.29414876 | CXCL10+ macrophage | No |
| EMP3 | 1.659891556 | 0.875 | 0.479 | 57.64382051 | CXCL10+ macrophage | Yes |
| ETV5 | -3.140533868 | 0.062 | 0.276 | 15.83397163 | SELENOP+ macrophage | No |
| ETV7 | 3.430054513 | 0.26 | 0.044 | 24.29622627 | CXCL10+ macrophage | No |
| F13A1 | -3.636694025 | 0.073 | 0.298 | 18.56945849 | SELENOP+ macrophage | No |
| FCGR1B | 2.05378696 | 0.523 | 0.21 | 28.55110322 | CXCL10+ macrophage | No |
| FCN1 | 2.375595232 | 0.905 | 0.248 | 117.329455 | CXCL10+ macrophage | No |
| FFAR2 | 3.815295723 | 0.271 | 0.019 | 34.59034864 | CXCL10+ macrophage | No |
| FGR | 2.287890952 | 0.696 | 0.236 | 59.07329892 | CXCL10+ macrophage | No |
| FLNA | 2.24298455 | 0.705 | 0.229 | 57.43410129 | CXCL10+ macrophage | No |
| FOLR2 | -2.798905993 | 0.141 | 0.357 | 18.01996902 | SELENOP+ macrophage | No |
| FPR1 | 1.65230723 | 0.547 | 0.292 | 21.96970884 | CXCL10+ macrophage | Yes |
| FPR2 | 3.286759548 | 0.19 | 0.026 | 15.51086328 | CXCL10+ macrophage | No |
| FRMD4A | -3.35966036 | 0.035 | 0.271 | 20.96902587 | SELENOP+ macrophage | No |
| FRMD4B | -2.401193025 | 0.173 | 0.396 | 21.53706695 | SELENOP+ macrophage | No |
| GAL3ST4 | -3.34765662 | 0.011 | 0.171 | 15.69294392 | SELENOP+ macrophage | No |
| GATM | -3.807146815 | 0.033 | 0.259 | 19.65801992 | SELENOP+ macrophage | No |
| GBP1 | 3.233267004 | 0.954 | 0.35 | 164.7504889 | CXCL10+ macrophage | Yes |
| GBP2 | 2.356651535 | 0.802 | 0.341 | 84.77209586 | CXCL10+ macrophage | Yes |
| GBP4 | 2.144697025 | 0.832 | 0.354 | 79.43509716 | CXCL10+ macrophage | Yes |
| GBP5 | 2.983100147 | 0.846 | 0.281 | 115.8519355 | CXCL10+ macrophage | Yes |
| GCH1 | 2.980373524 | 0.612 | 0.14 | 71.6525246 | CXCL10+ macrophage | No |
| GLIPR2 | 1.609617885 | 0.474 | 0.203 | 18.84535085 | CXCL10+ macrophage | No |
| GPBAR1 | 2.427141386 | 0.415 | 0.11 | 28.9433624 | CXCL10+ macrophage | No |

**SUPPLEMENTARY MATERIAL**

| | | | | | | |
|---|---|---|---|---|---|---|
| GPNMB | -4.61961238 | 0.03 | 0.271 | 23.43842618 | SELENOP+ macrophage | No |
| GPR34 | -5.529259836 | 0.019 | 0.397 | 48.72692398 | SELENOP+ macrophage | No |
| HAMP | -5.822672178 | 0.014 | 0.178 | 17.40415356 | SELENOP+ macrophage | No |
| HCK | 1.66670188 | 0.734 | 0.406 | 45.85844276 | CXCL10+ macrophage | No |
| HEXA | -2.053989981 | 0.184 | 0.43 | 17.19898325 | SELENOP+ macrophage | No |
| ICAM2 | 2.157371471 | 0.447 | 0.151 | 28.31698213 | CXCL10+ macrophage | Yes |
| IDO1 | 3.859606078 | 0.187 | 0.025 | 16.102267 | CXCL10+ macrophage | Yes |
| IFI35 | 1.910647267 | 0.588 | 0.247 | 35.47707344 | CXCL10+ macrophage | No |
| IFIT2 | 1.928435762 | 0.455 | 0.151 | 24.61927929 | CXCL10+ macrophage | No |
| IFIT3 | 2.511303748 | 0.512 | 0.128 | 43.52823808 | CXCL10+ macrophage | No |
| IFITM1 | 2.88827454 | 0.623 | 0.175 | 59.59310068 | CXCL10+ macrophage | Yes |
| IFITM2 | 1.963208264 | 0.799 | 0.528 | 61.47816922 | CXCL10+ macrophage | Yes |
| IFITM3 | 1.931109103 | 0.957 | 0.719 | 97.59384132 | CXCL10+ macrophage | Yes |
| IL18 | -2.370757445 | 0.214 | 0.495 | 29.2639837 | SELENOP+ macrophage | Yes |
| IRF1 | 1.789916608 | 0.848 | 0.452 | 57.10072073 | CXCL10+ macrophage | Yes |
| IRF7 | 1.750816283 | 0.612 | 0.276 | 33.21623294 | CXCL10+ macrophage | Yes |
| ISG20 | 2.179691719 | 0.444 | 0.125 | 31.75179832 | CXCL10+ macrophage | Yes |
| ITGAL | 1.678550215 | 0.485 | 0.222 | 20.33331183 | CXCL10+ macrophage | No |
| JUN | -2.76874089 | 0.198 | 0.506 | 27.28589459 | SELENOP+ macrophage | Yes |
| KYNU | 1.828379213 | 0.485 | 0.246 | 24.89590531 | CXCL10+ macrophage | No |
| LCP1 | 1.500373549 | 0.921 | 0.586 | 58.49070082 | CXCL10+ macrophage | No |
| LGALS2 | 1.601016255 | 0.455 | 0.187 | 16.26798052 | CXCL10+ macrophage | No |
| LGMN | -3.305283598 | 0.103 | 0.446 | 32.85464593 | SELENOP+ macrophage | No |
| LILRA1 | 2.592978603 | 0.425 | 0.084 | 37.58711906 | CXCL10+ macrophage | No |
| LILRA2 | 1.845588275 | 0.423 | 0.177 | 17.29466789 | CXCL10+ macrophage | No |
| LILRA5 | 2.897042423 | 0.531 | 0.104 | 54.95030082 | CXCL10+ macrophage | No |
| LILRB1 | 1.73820668 | 0.718 | 0.342 | 41.64961423 | CXCL10+ macrophage | Yes |
| LILRB2 | 1.908060894 | 0.762 | 0.361 | 55.97306746 | CXCL10+ macrophage | Yes |
| LRRK2 | 1.96744094 | 0.523 | 0.235 | 28.07009911 | CXCL10+ macrophage | No |
| LST1 | 1.629173741 | 0.911 | 0.715 | 77.25546694 | CXCL10+ macrophage | Yes |
| LTC4S | -4.824090709 | 0.027 | 0.323 | 32.63893747 | SELENOP+ macrophage | No |
| LY6E | 2.411357293 | 0.829 | 0.325 | 87.71271086 | CXCL10+ macrophage | No |
| LYN | 1.555931271 | 0.694 | 0.404 | 37.07839798 | CXCL10+ macrophage | No |
| LYST | 2.018204309 | 0.523 | 0.221 | 31.71465621 | CXCL10+ macrophage | No |
| MGAT4A | -3.357649715 | 0.057 | 0.366 | 29.30556404 | SELENOP+ macrophage | No |
| MS4A4A | -2.396589913 | 0.195 | 0.456 | 23.45247334 | SELENOP+ macrophage | Yes |
| MYD88 | 1.697608689 | 0.572 | 0.29 | 30.07044562 | CXCL10+ macrophage | Yes |
| MYO1G | 3.327511741 | 0.491 | 0.076 | 57.44314147 | CXCL10+ macrophage | No |
| MYOF | 1.745244503 | 0.604 | 0.271 | 29.04749184 | CXCL10+ macrophage | No |
| NBN | 1.64741162 | 0.409 | 0.192 | 15.73717366 | CXCL10+ macrophage | No |
| NCF1 | 1.559903591 | 0.767 | 0.512 | 31.71354336 | CXCL10+ macrophage | No |
| NEDD9 | 2.094163879 | 0.287 | 0.063 | 20.69974894 | CXCL10+ macrophage | No |

**SUPPLEMENTARY MATERIAL**

| | | | | | | |
|---|---|---|---|---|---|---|
| NR4A2 | -2.43458311 | 0.13 | 0.361 | 15.82067553 | SELENOP+ macrophage | No |
| NRP1 | -4.37395202 | 0.019 | 0.209 | 16.69706066 | SELENOP+ macrophage | No |
| OAS3 | 2.194346222 | 0.336 | 0.104 | 18.29423338 | CXCL10+ macrophage | No |
| OLFML3 | -4.116968743 | 0.024 | 0.208 | 15.37893249 | SELENOP+ macrophage | No |
| OLR1 | -3.925129078 | 0.041 | 0.331 | 29.4341435 | SELENOP+ macrophage | No |
| PARP14 | 1.504567923 | 0.699 | 0.395 | 32.71540618 | CXCL10+ macrophage | No |
| PEBP1 | -1.913709161 | 0.249 | 0.506 | 20.58256114 | SELENOP+ macrophage | No |
| PLAC8 | 2.624086125 | 0.534 | 0.144 | 47.61180479 | CXCL10+ macrophage | No |
| PLD3 | -1.900837103 | 0.322 | 0.521 | 22.3390861 | SELENOP+ macrophage | No |
| PLD4 | -3.172721707 | 0.076 | 0.286 | 17.66236126 | SELENOP+ macrophage | No |
| PLEK | 2.188471342 | 0.864 | 0.498 | 79.82327222 | CXCL10+ macrophage | No |
| PLTP | -5.035833895 | 0.043 | 0.346 | 33.65339376 | SELENOP+ macrophage | No |
| PSMB9 | 1.584929894 | 0.889 | 0.547 | 67.84365305 | CXCL10+ macrophage | Yes |
| PSTPIP2 | 1.906420309 | 0.621 | 0.255 | 36.87616922 | CXCL10+ macrophage | No |
| PTGIR | 2.565827251 | 0.252 | 0.033 | 24.64054385 | CXCL10+ macrophage | No |
| PTPN6 | 1.567782776 | 0.751 | 0.447 | 38.75463354 | CXCL10+ macrophage | Yes |
| RAB24 | 2.271073286 | 0.58 | 0.216 | 49.64917695 | CXCL10+ macrophage | No |
| RGS1 | -3.757433892 | 0.049 | 0.33 | 26.19864541 | SELENOP+ macrophage | No |
| RIPOR2 | 2.994304764 | 0.434 | 0.081 | 42.35296917 | CXCL10+ macrophage | No |
| RNASE1 | -5.394587359 | 0.041 | 0.283 | 23.90458671 | SELENOP+ macrophage | No |
| RNASE6 | -2.077007199 | 0.179 | 0.497 | 23.75281782 | SELENOP+ macrophage | No |
| RNF144B | 1.829260614 | 0.512 | 0.274 | 22.57911835 | CXCL10+ macrophage | No |
| S100A4 | 1.741946603 | 0.935 | 0.66 | 61.7596719 | CXCL10+ macrophage | No |
| S100A6 | 1.844705604 | 0.908 | 0.62 | 58.54577073 | CXCL10+ macrophage | No |
| S100A8 | 2.37445842 | 0.539 | 0.153 | 38.82103232 | CXCL10+ macrophage | Yes |
| S100A9 | 2.210258686 | 0.715 | 0.271 | 48.4935087 | CXCL10+ macrophage | Yes |
| SAMD9L | 1.808411806 | 0.512 | 0.23 | 23.51354037 | CXCL10+ macrophage | No |
| SCIMP | 1.587868115 | 0.566 | 0.276 | 24.95439855 | CXCL10+ macrophage | No |
| SECTM1 | 2.454832707 | 0.36 | 0.1 | 23.89029644 | CXCL10+ macrophage | No |
| SELENOP | -7.327089965 | 0.024 | 0.368 | 45.75605598 | SELENOP+ macrophage | No |
| SEPTIN9 | 1.970292321 | 0.42 | 0.173 | 22.13150531 | CXCL10+ macrophage | No |
| SERPINB9 | 1.591519955 | 0.583 | 0.279 | 24.54322729 | CXCL10+ macrophage | No |
| SERPINF1 | -3.834955473 | 0.027 | 0.28 | 24.18845731 | SELENOP+ macrophage | No |
| SGK1 | -2.099081531 | 0.233 | 0.487 | 18.66208558 | SELENOP+ macrophage | No |
| SLAMF7 | 2.925834 | 0.631 | 0.174 | 70.30810172 | CXCL10+ macrophage | Yes |
| SLC2A6 | 3.307871739 | 0.453 | 0.087 | 50.14810951 | CXCL10+ macrophage | No |
| SLC31A2 | 1.640226415 | 0.583 | 0.306 | 26.93178713 | CXCL10+ macrophage | No |
| SLC40A1 | -3.988779022 | 0.084 | 0.381 | 30.71702383 | SELENOP+ macrophage | No |
| SLCO2B1 | -3.988222417 | 0.06 | 0.438 | 43.64004234 | SELENOP+ macrophage | No |
| SMIM25 | 3.509174742 | 0.661 | 0.154 | 87.61760912 | CXCL10+ macrophage | No |
| SOD2 | 1.898784017 | 0.916 | 0.553 | 70.3996555 | CXCL10+ macrophage | Yes |
| SP110 | 1.678022114 | 0.637 | 0.311 | 30.76457408 | CXCL10+ macrophage | No |

| | | | | | | |
|---|---|---|---|---|---|---|
| STAB1 | -2.757873687 | 0.163 | 0.381 | 21.18190803 | SELENOP+ macrophage | No |
| STEAP4 | 4.874277373 | 0.146 | 0.007 | 18.27073807 | CXCL10+ macrophage | No |
| STX11 | 1.654631459 | 0.561 | 0.229 | 29.19030783 | CXCL10+ macrophage | No |
| STXBP2 | 1.517245113 | 0.675 | 0.367 | 27.34562828 | CXCL10+ macrophage | No |
| TAP1 | 1.95972257 | 0.713 | 0.312 | 50.73036437 | CXCL10+ macrophage | Yes |
| TCF7L2 | 2.513919072 | 0.523 | 0.145 | 43.00882054 | CXCL10+ macrophage | No |
| TM6SF1 | -2.95902856 | 0.049 | 0.27 | 16.33902914 | SELENOP+ macrophage | No |
| TMEM131L | 2.430332612 | 0.382 | 0.135 | 25.90808501 | CXCL10+ macrophage | No |
| TNFAIP2 | 1.975625312 | 0.805 | 0.384 | 64.59839249 | CXCL10+ macrophage | No |
| TNFSF10 | 2.982181527 | 0.848 | 0.308 | 120.2292742 | CXCL10+ macrophage | Yes |
| TREM2 | -5.255512195 | 0.03 | 0.391 | 45.39030119 | SELENOP+ macrophage | No |
| VAMP5 | 1.577258337 | 0.889 | 0.517 | 55.82088305 | CXCL10+ macrophage | No |
| VASP | 1.691637129 | 0.696 | 0.346 | 40.2120235 | CXCL10+ macrophage | No |
| VCAN | 1.751322207 | 0.309 | 0.085 | 15.96217033 | CXCL10+ macrophage | Yes |
| VSIG4 | -2.6501941 | 0.084 | 0.352 | 20.57900983 | SELENOP+ macrophage | No |
| WARS | 3.194709592 | 0.954 | 0.366 | 156.0411257 | CXCL10+ macrophage | Yes |
| YWHAH | -2.146768934 | 0.255 | 0.506 | 23.79678401 | SELENOP+ macrophage | No |

Table S3: *CXCL10*+ and *SELENOP*+ macrophages gene signatures in scRNAseq. The pct columns represent the proportion of cells (from 0 to 1) in each macrophage subset expressing the indicated gene.
